# The Gastric Cancer Registry: A Genomic Translational Resource for Multidisciplinary Research in Stomach Malignancies

**DOI:** 10.1101/2022.03.10.22271914

**Authors:** Alison F Almeda, Sue Grimes, HoJoon Lee, Stephanie U Greer, GiWon Shin, Madeline McNamara, Anna C Hooker, Maya Arce, Matthew Kubit, Marie Schauer, Paul Van Hummelen, Cindy Ma, Meredith Mills, Robert J Huang, Joo Ha Hwang, Manuel R Amieva, Summer Han, James M. Ford, Hanlee P. Ji

**Affiliations:** Division of Oncology, Department of Medicine, Stanford University School of Medicine, Stanford, CA, 94305, United States; Division of Gastroenterology and Hepatology, Department of Medicine, Stanford University School of Medicine, Stanford, CA, 94305, United States; Division of Infectious Diseases, Department of Pediatrics, Stanford University School of Medicine, Stanford, CA, 94305, United States; Department of Neurosurgery, Stanford University School of Medicine, Stanford, CA, 94305, United States

**Keywords:** Gastric Cancer, Cancer Registry, Hereditary Cancer, Sequencing, Data Portal

## Abstract

**Background:** Gastric cancer **(GC)** is a leading cause of global cancer morbidity and mortality. Developing information systems which integrate clinical and genomic data may accelerate discoveries to improve cancer prevention, detection, and treatment. To support translational research in GC, we developed the GC Registry **(GCR)**, a North American repository of clinical and cancer genomics data.

**Methods:** GCR is a national registry with online self-enrollment. Entry criteria into the GCR included the following: (1) diagnosis of GC, (2) history of GC in a first-or second-degree family member or (3) known pathogenic or likely pathogenic germline mutation in the gene *CDH1*. Participants provided demographic and clinical information through a detailed (412-item) online survey. A subset of participants provided specimens of saliva and tumor samples. These tumor samples underwent exome sequencing, whole genome sequencing and transcriptome sequencing.

**Results:** From 2011-2021, 567 individuals registered for the GCR and returned the clinical questionnaire. For this cohort 65% had a personal history of GC, 36% reported a family history of GC and 14% had a germline *CDH1* mutation. Eighty-nine GC patients provided tumor tissue samples. For the initial pilot study, 41 tumors were sequenced using next generation sequencing. The data was analyzed for cancer mutations, copy number variations, gene expression, microbiome presence, neoantigens, immune infiltrates, and other features. We developed a searchable, web-based interface (the GCR Genome Explorer) to enable researchers access to these datasets.

**Conclusions:** The GCR is a unique, North American GC registry which integrates both clinical and genomic annotation.

**Impact:** Available for researchers through an open access, web-based explorer, we hope the GCR Genome Explorer accelerates collaborative GC research across the United States.

## INTRODUCTION

Gastric cancer **(GC)** is a leading cause of cancer morbidity and mortality worldwide.^1^ While incidence is lower in the United States, GC remains a major public health concern with an estimated 116,000 prevalent cases nationwide in 2017.^2^ GC is diagnosed at generally advanced stages in the United States, where curative resection is often no longer possible.^2,3^ These data underscore the need for additional translational research in GC etiology, prevention, early detection, and therapy.

Cancer registries are a valuable resource for collating clinical information. Some registries also contain biological specimen repositories of both cancerous and non-cancerous tissue, allowing for somatic and germline genomic characterization through next generation sequencing.^4,5^ Cancer registries that integrate clinical information with tumor genomic features are particularly useful in translational research.^6,7^ There currently exist few registries focused on gastric cancer, particularly with patient data and samples derived from the United States. The availability of medical records, epidemiological data, and biospecimens of tumor tissue allow researchers to analyze genetic, environmental, and other differences that provide clues leading to risk factors. This research is particularly relevant given the poor overall outcomes from GC in the United States, and lack of established screening and surveillance programs for this deadly cancer.

To address this knowledge gap, we established the Gastric Cancer Registry **(GCR)** in 2011. The goal of this project is to integrate granular patient clinical data (collected through a detailed, 412-item online questionnaire) with comprehensive genomic characterization of tumor samples. This includes gene expression, somatic mutations, copy number variation, human leukocyte antigen genotypes, neoantigens, and intra-tumoral heterogeneity details. To facilitate public access to these data, we created the GCR Genome Explorer (https://gcregistry-explorer.stanford.edu/), a browser-based interactive tool which allows for querying of clinical and molecular annotation from the GCR. In this manuscript, we describe the overall study design, methods for sample collection and data generation, and characteristics of enrolled participants in the GCR over a ten-year period (2011-2021). We also review features of the GCR Genome Explorer and describe how this tool can be used by cancer researchers for translational research.

## MATERIALS AND METHODS

### Study design

Approval was obtained from the Stanford University Institutional Review Board (IRB-20285). The study was performed in accordance with the ethical standards delineated in the 1964 Declaration of Helsinki and its later amendments. Eligible participants were over 18 years of age and fulfilled one or more of the following criteria: personal history of histologically proven GC, a family history of GC in a first-or second-degree relative, and/or a known pathogenic or likely pathogenic mutation in the gene *CDH1*, also known as Cadherin-1. From 2011-2014, only individuals with a GC diagnosis were included in the study. Family history and a known germline *CDH1* mutation were added as eligibility criteria in 2014. Biospecimens were collected from residents of the United States. Recruitment and enrollment for the GCR was conducted through referral by local clinicians, local and regional pamphlet distribution at conferences and interest/advocacy group events, social media advertisement and a study website (https://gcregistry.stanford.edu/). Registrants use the website to complete the questionnaire.

Study data were collected and managed using REDCap electronic data capture tools hosted at Stanford University. REDCap is a secure, web-based software platform designed to support data capture for research studies, providing 1) an intuitive interface for validated data capture; 2) audit trails for tracking data manipulation and export procedures; 3) automated export procedures for seamless data downloads to common statistical packages; and 4) procedures for data integration and interoperability with external sources.^8,9^ Moreover, REDCap allows for automated consent for participation in research. Upon registration, participants were asked to complete a 412-item questionnaire (**Supplementary File 1**). This intake survey is focused on information relevant to genetics, lifestyle, environmental and personal risk factors related to GC.

The questionnaire contained questions regarding subjects’ demographics, medical history, familial cancer history, and lifestyle behaviors. For providing updates in personal or family history, participants could provide additional information using a unique return code. Participants had the option of authorizing the release of medical records related to the diagnosis and treatment of the patient’s GC. This information included treatment summaries from any chemotherapy or radiation therapy, the operative notes from any surgeries and any pathology reports relating to the diagnosis of GC.

### Biospecimen collection

We obtained tissue specimens for a subset of participants. These samples included archival tissue samples in the form of formalin fixed, paraffin embedded **(FFPE)** blocks, scrolls and unstained slides. This cohort included individuals with GC who had a surgical resection of the stomach or carriers of *CDH-1* germline mutations who underwent a screening endoscopic with biopsy or preventative gastrectomy. From 2018 onward, participants had the option of donating a sample of saliva. We used the Oragene DNA collection kit (DNA Genotek Inc., catalog no. OGR-500, OGR-600) to collect these samples.

### Sample processing

Archival FFPE samples of gastric tumors were used for genomic studies. Genomic DNA and total RNA were extracted from the tumor samples and the quality of nucleic acids was determined through quantitation, genomic sizing, and fragmentation analysis. We used an ultrasonicator to shear genomic DNA to a desired length of 300 base pairs. The genomic DNA was transformed into sequencing libraries using the KAPA Hyper Prep Kit for Illumina (Roche, catalog no. KK8502) with eight-base pair unique dual adapters. An amount of genomic DNA library was set aside for exome enrichment using xGen Lockdown Probes and reagents (Integrated DNA Technologies, catalog no. 1056114, 1075474). To prepare RNA sequencing libraries, the KAPA RNA HyperPrep Kit (Roche, catalog no. KK8540) was used with modifications to accommodate low quality samples. The genomic DNA, exome, and RNA libraries were pooled respectively and sequenced on the NovaSeq 6000 system (Illumina, catalog no. 20012850). More details about the methods are described in **Supplementary File 2**.

### Next generation sequencing

The genomic DNA libraries underwent low-coverage 1-2X whole genome sequencing. Exome libraries were sequenced more deeply at approximately 68X coverage to enable somatic mutation calling. The RNA libraries were sequenced at high depth for an average of 65 million reads per sample.

### Bioinformatic analysis

Whole genome sequencing reads were analyzed for copy number variation. To identify somatic copy number changes for samples without a matched normal control, we used a normal reference genome data set as a comparison control.^10^ Whole exome sequencing data was analyzed for single nucleotide variants and insertions/deletions of nucleotides in genomic DNA. RNA-seq data was investigated for multiple features, including: (i) gene expression level, (ii) four-digit human leukocyte antigen (**HLA**) genotypes, (iii) tumor immune infiltrate cell types, and (iv) the microbiome. Based on whole exome sequencing data and RNA-seq, candidate cancer neoantigens were identified. More detailed methods and bioinformatic pipeline specifics are in **Supplementary File 2**.

### The GCR Genome Explorer

The GCR Genome Explorer contains clinical, genomic, and genetic data from the registry. Within the GCR Genome Explorer, we also included other datasets derived from the Cancer Genome Atlas **(TCGA)** stomach adenocarcinoma **(STAD)** and esophageal carcinoma **(ESCA)** projects. The Variant Call Format files and copy number variations for the TCGA cohorts were downloaded from the National Cancer Institute Genomic Data Commons.^11^ Genomic and genetic TCGA data was processed using the similar bioinformatics pipeline as GCR data (though with changes to accommodate for matched tumor-normal data), resulting in comparable bioinformatic analysis across all samples.

The portal is a two-tier client-server application written using Ruby on Rails version 5.1.7 and ruby 2.4.9, with back-end database tables in MySQL 5.5.62 and deployed using Passenger and Apache2. The application server has 64GB RAM and 32 processors running Ubuntu 16.04. The database server has 32GB RAM and 16 processors running Ubuntu 16.04.

The user interface utilizes bootstrap version 3.4.1 for responsive sizing to different format clients and browsers. Standard formatting, search, and filtering capability for query tables is provided by the jQuery DataTables plugin. Highcharts is used for generation of all plots. All queries and plots are produced dynamically from the underlying database tables based on user query parameters. The URL is https://gcregistry-explorer.stanford.edu/.

## RESULTS

### Participant population and demographics

From March 2011 to November 2021, 567 subjects enrolled in the study. For inclusion in the study, all participants were required to identify their eligibility status on the enrollment questionnaire. The majority reported a personal history of GC. Some participants met multiple eligibility criteria (*e*.*g*., having GC and a family history of GC). Eligibility status with respect to sex, age, race, and ethnicity is depicted in **Table 1**. Participants were predominantly female (63%), White (76%), and non-Hispanic (53%). The median age of all participants was 51 years (range: 18-92 years). The median age of participants with GC was 68 years. Some participants did not report their sex, race, ethnicity, or other demographic details as these questions were optional on the enrollment survey.

**Table 1.**
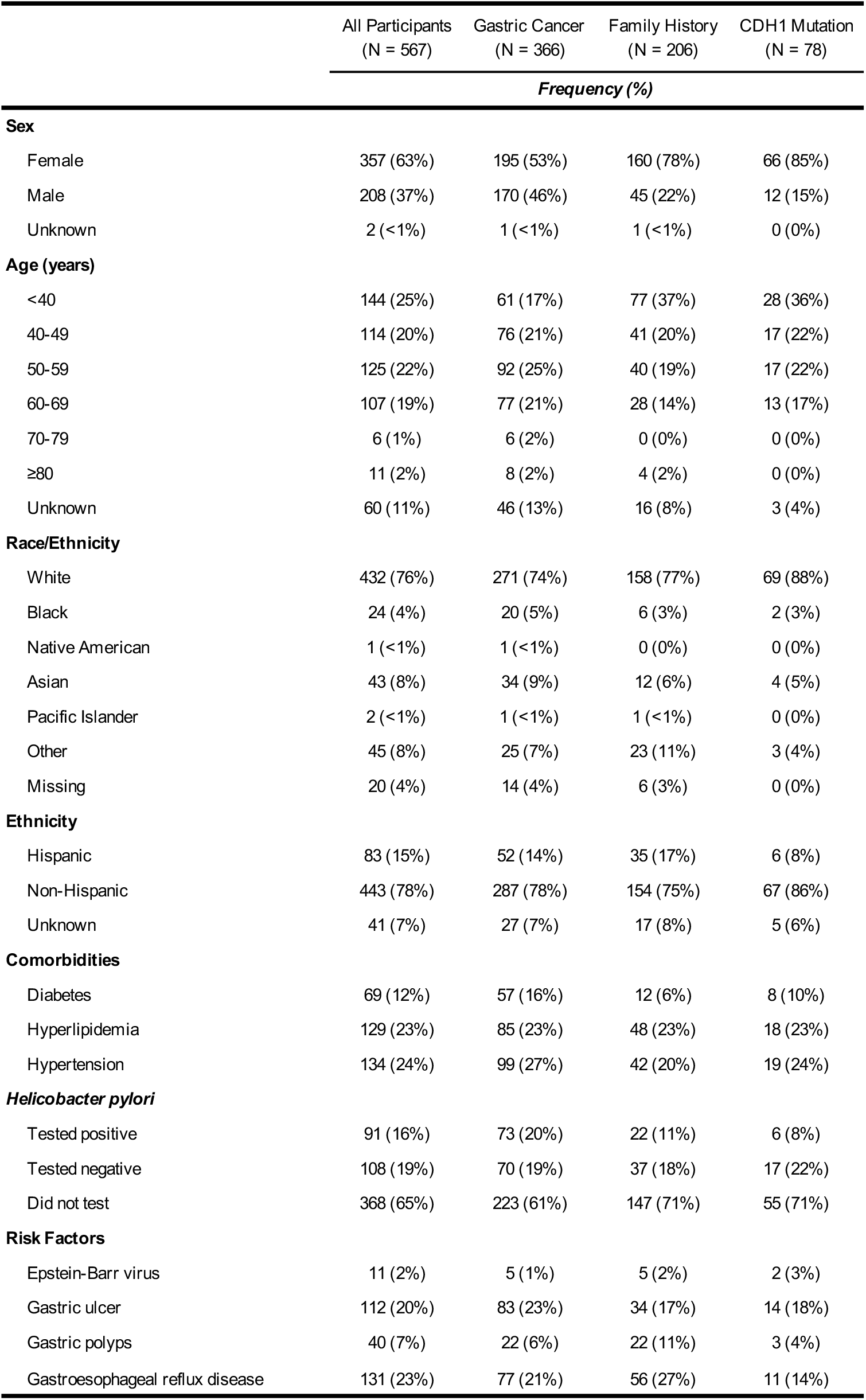
Cohort characteristics by eligibility status from 2011-2021. As participants could fall into more than one eligibility category, some participants appear in multiple columns.

|  | All Participants<br>(N = 567) | Gastric Cancer<br>(N = 366) | Family History<br>(N = 206) | CDH1 Mutation<br>(N = 78) |
| --- | --- | --- | --- | --- |
| Frequency (%) |  |  |  |  |
| Sex |  |  |  |  |
| Female | 357 (63%) | 195 (53%) | 160 (78%) | 66 (85%) |
| Male | 208 (37%) | 170 (46%) | 45 (22%) | 12 (15%) |
| Unknown | 2 (<1%) | 1 (<1%) | 1 (<1%) | 0 (0%) |
| Age (years) |  |  |  |  |
| <40 | 144 (25%) | 61 (17%) | 77 (37%) | 28 (36%) |
| 40-49 | 114 (20%) | 76 (21%) | 41 (20%) | 17 (22%) |
| 50-59 | 125 (22%) | 92 (25%) | 40 (19%) | 17 (22%) |
| 60-69 | 107 (19%) | 77 (21%) | 28 (14%) | 13 (17%) |
| 70-79 | 6 (1%) | 6 (2%) | 0 (0%) | 0 (0%) |
| ≥80 | 11 (2%) | 8 (2%) | 4 (2%) | 0 (0%) |
| Unknown | 60 (11%) | 46 (13%) | 16 (8%) | 3 (4%) |
| Race/Ethnicity |  |  |  |  |
| White | 432 (76%) | 271 (74%) | 158 (77%) | 69 (88%) |
| Black | 24 (4%) | 20 (5%) | 6 (3%) | 2 (3%) |
| Native American | 1 (<1%) | 1 (<1%) | 0 (0%) | 0 (0%) |
| Asian | 43 (8%) | 34 (9%) | 12 (6%) | 4 (5%) |
| Pacific Islander | 2 (<1%) | 1 (<1%) | 1 (<1%) | 0 (0%) |
| Other | 45 (8%) | 25 (7%) | 23 (11%) | 3 (4%) |
| Missing | 20 (4%) | 14 (4%) | 6 (3%) | 0 (0%) |
| Ethnicity |  |  |  |  |
| Hispanic | 83 (15%) | 52 (14%) | 35 (17%) | 6 (8%) |
| Non-Hispanic | 443 (78%) | 287 (78%) | 154 (75%) | 67 (86%) |
| Unknown | 41 (7%) | 27 (7%) | 17 (8%) | 5 (6%) |
| Comorbidities |  |  |  |  |
| Diabetes | 69 (12%) | 57 (16%) | 12 (6%) | 8 (10%) |
| Hyperlipidemia | 129 (23%) | 85 (23%) | 48 (23%) | 18 (23%) |
| Hypertension | 134 (24%) | 99 (27%) | 42 (20%) | 19 (24%) |
| Helicobacter pylori |  |  |  |  |
| Tested positive | 91 (16%) | 73 (20%) | 22 (11%) | 6 (8%) |
| Tested negative | 108 (19%) | 70 (19%) | 37 (18%) | 17 (22%) |
| Did not test | 368 (65%) | 223 (61%) | 147 (71%) | 55 (71%) |
| Risk Factors |  |  |  |  |
| Epstein-Barr virus | 11 (2%) | 5 (1%) | 5 (2%) | 2 (3%) |
| Gastric ulcer | 112 (20%) | 83 (23%) | 34 (17%) | 14 (18%) |
| Gastric polyps | 40 (7%) | 22 (6%) | 22 (11%) | 3 (4%) |
| Gastroesophageal reflux disease | 131 (23%) | 77 (21%) | 56 (27%) | 11 (14%) |

The participants’ commonly reported medications included non-steroidal anti-inflammatories, proton pump inhibitors and multivitamins. Common comorbidities included high blood pressure, high cholesterol, and other cancers. Nearly a quarter of participants with GC reported a history of gastric ulcers, gastroesophageal reflux disease, or gastric polyps. *Helicobacter pylori* infection was reported by 16% of all participants, and 20% of all participants with GC. Notably, a high proportion of participants (65%) did not report or did not know their *H. pylori* status.

### Biospecimens

For this study,164 participants donated biological specimens. We collected 111 saliva samples and 89 tissue samples. At the time of publication, 41 tumor GCR samples had undergone sequencing and with the results available on the GCR Genome Explorer **(Table 2)**. This subset of GCR archival samples underwent histopathological review, and most patients were diagnosed with gastric adenocarcinoma. Clinical and histologic characteristics of the sequenced tumors are depicted in **Table 2**. Of the specimens where Lauren’s classification was reported (N=24), the majority were diffuse-type cancers (N=16). The tumors were generally aggressive and poorly differentiated (59%). The majority of tumors were from patients with metastatic disease (54%). With respect to anatomic location, 10% of tumors arose from the cardia, 73% arose from the non-cardia stomach, and 17% arose from an unreported location.

**Table 2.** Overview of gastric tumor tissues in the GCR Genome Explorer (N=41).

|  | Frequency (%) |
| --- | --- |
| **Sex** |  |
| Male | 24 (59%) |
| Female | 16 (39%) |
| Not available | 1 (2%) |
| **Histology** |  |
| Gastric adenocarcinoma | 37 (90%) |
| GIST | 1 (2%) |
| Not available | 2 (5%) |
| **Histological Subtype** |  |
| Intestinal | 4 (10%) |
| Diffuse | 16 (39%) |
| Mixed | 4 (10%) |
| Not available | 17 (41%) |
| **Histological differentiation** |  |
| Well | 1 (2%) |
| Moderate | 3 (7%) |
| Moderately to poorly | 3 (7%) |
| Poorly | 24 (59%) |
| Not available | 10 (24%) |
| **AJCC tumor pathology** |  |
| T1 | 6 (15%) |
| T2 | 0 (0%) |
| T3 | 12 (29%) |
| T4 | 7 (17%) |
| Not available | 16 (39%) |
| N0 | 9 (22%) |
| N1 | 3 (7%) |
| N2 | 4 (10%) |
| N3 | 7 (17%) |
| Not available | 18 (44%) |
| M0 | 1 (2%) |
| MX | 22 (54%) |
| Not available | 18 (44%) |
| **Lymph node involvement** |  |
| Positive | 13 (32%) |
| Negative | 12 (29%) |
| Not available | 16 (39%) |
| **Adjuvant radiation** |  |
| Yes | 14 (34%) |
| No | 20 (49%) |
| Not available | 7 (17%) |
| **Tumor anatomic site** |  |
| Gastroesophageal junction | 5 (12%) |
| Cardia | 4 (10%) |
| Pylorus | 2 (5%) |
| Fundus | 3 (7%) |
| Body | 8 (20%) |
| Antrum | 5 (12%) |
| Entire stomach | 7 (17%) |
| Not available | 7 (17%) |

### The GCR Genome Explorer and pilot data release

Public access is to the GCR Genome Explorer is available upon registration via the following URL: https://gcregistry-explorer.stanford.edu/users/sign_in. At time of publication, the Genome Explorer contains data from the initial 41 sequenced tumors through the GCR. In addition, genomic data from 443 TCGA gastric cancers and 185 TCGA esophageal cancers are available through the GCR Genome Explorer for cross-reference. Future releases will incorporate data from additional GCR tumor samples. A representative image of the GCR Genome Explorer home page showing available data sets is depicted in **Figure 1**.

**Figure 1.**
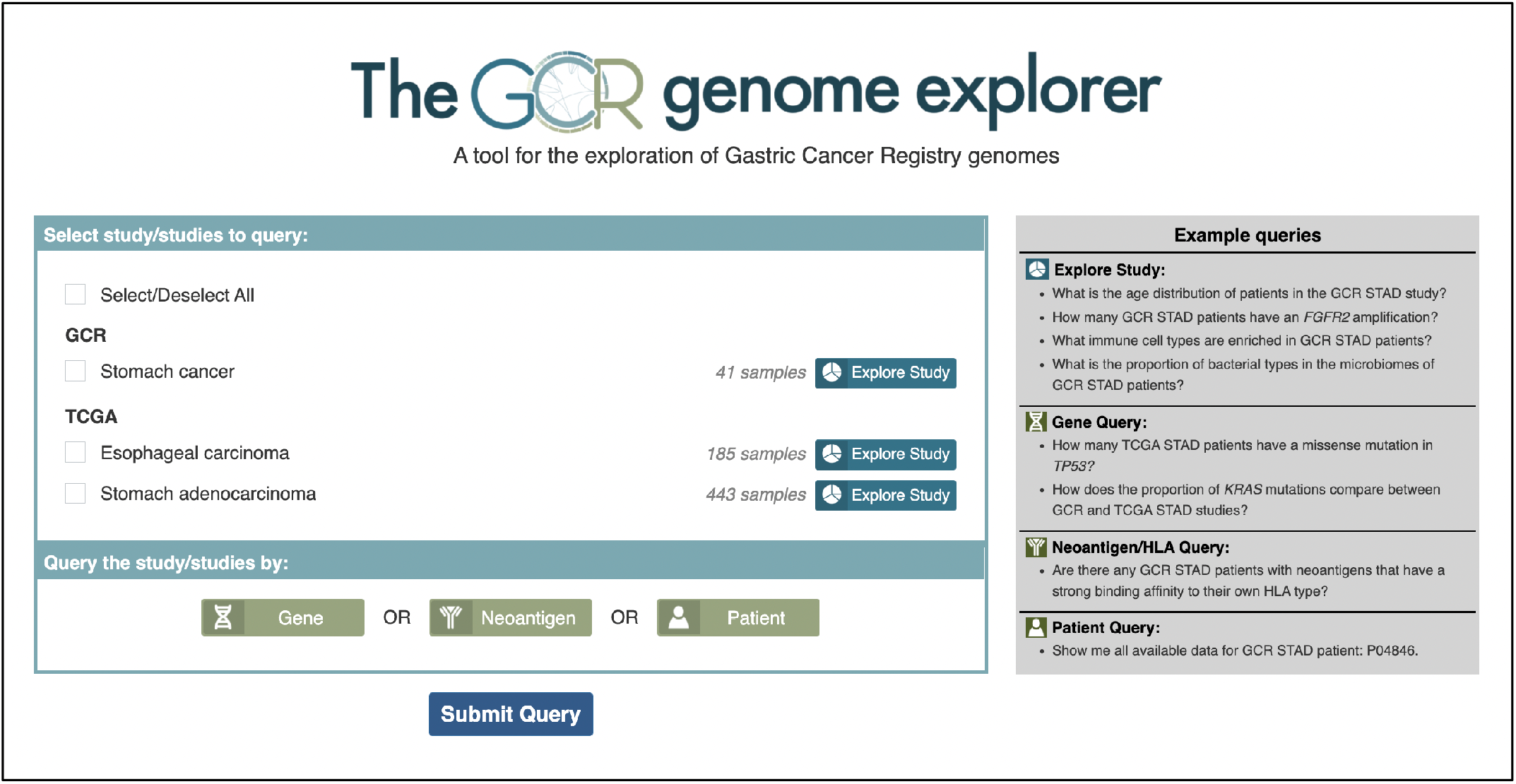
The GCR Genome Explorer home page. The portal offers queries of gene, neoantigen and patient datasets from one or multiple cohorts: the GC Registry (GCR), the Cancer Genome Atlas (TCGA) esophageal carcinoma (ESCA) study, and the TCGA stomach adenocarcinoma (STAD) study. The “Explore Study” feature enables the user to see the clinical and genomic characteristics of a single cohort.

There are two tiers of results that are provided in the Genome Explorer. The first tier includes gene expression levels determined from RNA-seq, somatic copy number based on whole genome sequencing and somatic mutations derived from exome sequencing. The second tier leverages the first-tier results - different algorithms extrapolate characteristics of the clonal diversity (WGS, exome), cellular microenvironment (RNA-seq), microbiome content (RNA-seq), HLA genotypes (RNA-seq) and putative neoantigens (WGS, exome, RNA-seq).

Cellular and microbiome features reflect the content of the local tumor microenvironment. We extrapolated the cellular representation and microbial populations using each tumor’s RNA-seq data. The cell results were based on processing with a deconvolution tool called CIBERSORTx.^12^ The analysis approximated the different cell types present in the tumor microenvironment.^13^ For those RNA-seq reads which did not align to the human genome, we used the Kraken2 program to determine if there were microbiome features that included bacterial genera.^14^

Nonsynonymous mutations are a source of immunogenic peptides, called neoantigens, which are tumor-specific and not expressed in other normal cells. Tumor mutational load, and more specifically, neoantigen load have been correlated with extent of T-cell reactivity, response to checkpoint therapy, and prognosis.^15-17^ Exome sequencing of these tumors allowed us to identify nonsynonymous mutations in the protein-coding portions of genes in cancer patients and predict potential neoantigens. We identified candidate neoantigens from the exome and RNA-seq data **(Methods)**. A candidate neoantigen fulfilled the following criteria: i) nonsynonymous somatic mutation, ii) expressed in transcriptome data and iii) translated neopeptides with strong binding affinity to patient’s own major histocompatibility molecules. Using a combination of the exome and transcriptome data and major histocompatibility complex genotypes, we generated a list of potential neoantigens for each tumor.

### GCR Genome Explorer features

There are several ways of accessing results through the GCR Genome Explorer. Options include general summaries of the results as well as specific queries. All image files and tables are available for download. On the home page (**Figure 1**), the different sample sets are displayed. For example, a user can query the GCR data set using the “Explore Study” feature, which directs to a landing page with multiple tabs. In the “Clinical Parameters” tab, the user will find visual and tabular representations of cohort characteristics. The cohort can be queried with regards to sex, race, ethnicity, cancer site, age at diagnosis, body mass index, smoking history, cancer diagnosis, Lauren classification, and histologic differentiation. Under “Gene Summary,” there are two tables describing genes that are most frequently mutated or varied in copy number within the GCR cohort. All cancer-associated genes are listed, along with genes mutated in >10% of samples, or genes with copy number variation in >10% of samples. The percentage filter will change across cohorts due to rounding, and due to gene ranking ties (i.e., multiple genes being mutated across the same total number of samples). The tables include the gene name, cytoband, number of samples displaying the mutation or copy number variation and ranked order of genes based on the percentage of samples affected. In addition, we provide annotations for genes that are cancer-associated, oncogenes or tumor-suppressor genes. Copy number summaries of the pilot data set showed that many tumor samples had a high degree of genomic instability. Whole genome sequencing revealed extensive changes in gene copy number, with some amplified genes such as *ERBB2* (i.e., *HER2*) being clinically actionable. Amplifications of *ERBB2* are an indication for the use of trastuzumab, a therapeutic monoclonal antibody. Exome sequencing allowed us to detect specific types of mutations. Each tumor sample contained unique combinations of missense mutations, frame shift deletions, and in-frame deletions across various genes.

The tab “HLA Types” contains an overview and breakdown of specific HLA alleles for 20 patients in the GCR study. There are options to view the prevalence of specific HLA-A, HLA-B, and HLA-C types within the study and within individual patients. In the next tab, “Immune Cells,” tumor-infiltrating immune cell types are identified and quantified in a bar plot as a percentage of overall immune cells present within a patient’s tumor. Alternatively, users can view the immune cell presence represented as a heatmap. The last tab, “Microbiome,” contains an interactive bar chart displaying the major bacteria phylum found within each patient. Users can select and deselect phylum of interest for a more granular, sample-specific view of the microbiome composition.

For more discrete summaries, users can employ the “Gene” query function to view data for a specific gene. We cite an example using the *CDH1* tumor suppressor gene. When searching all studies for *CDH1* gene information, the user will find discrete percentages and counts on its mutations, copy number variations, expression levels, and neoantigens within individual patients and across all cohorts (GCR and TCGA, **Figure 2**). The *CDH1* gene was mutated in 15.4% of GCR samples, 11% of TCGA-STAD samples and 1.1% of TCGA-ESCA samples. Nearly all samples in the GCR, TCGA-STAD and TCGA-ESCA cohorts showed high expression of *CDH1* (100%, 99.5%, and 99.4%).

**Figure 2.**
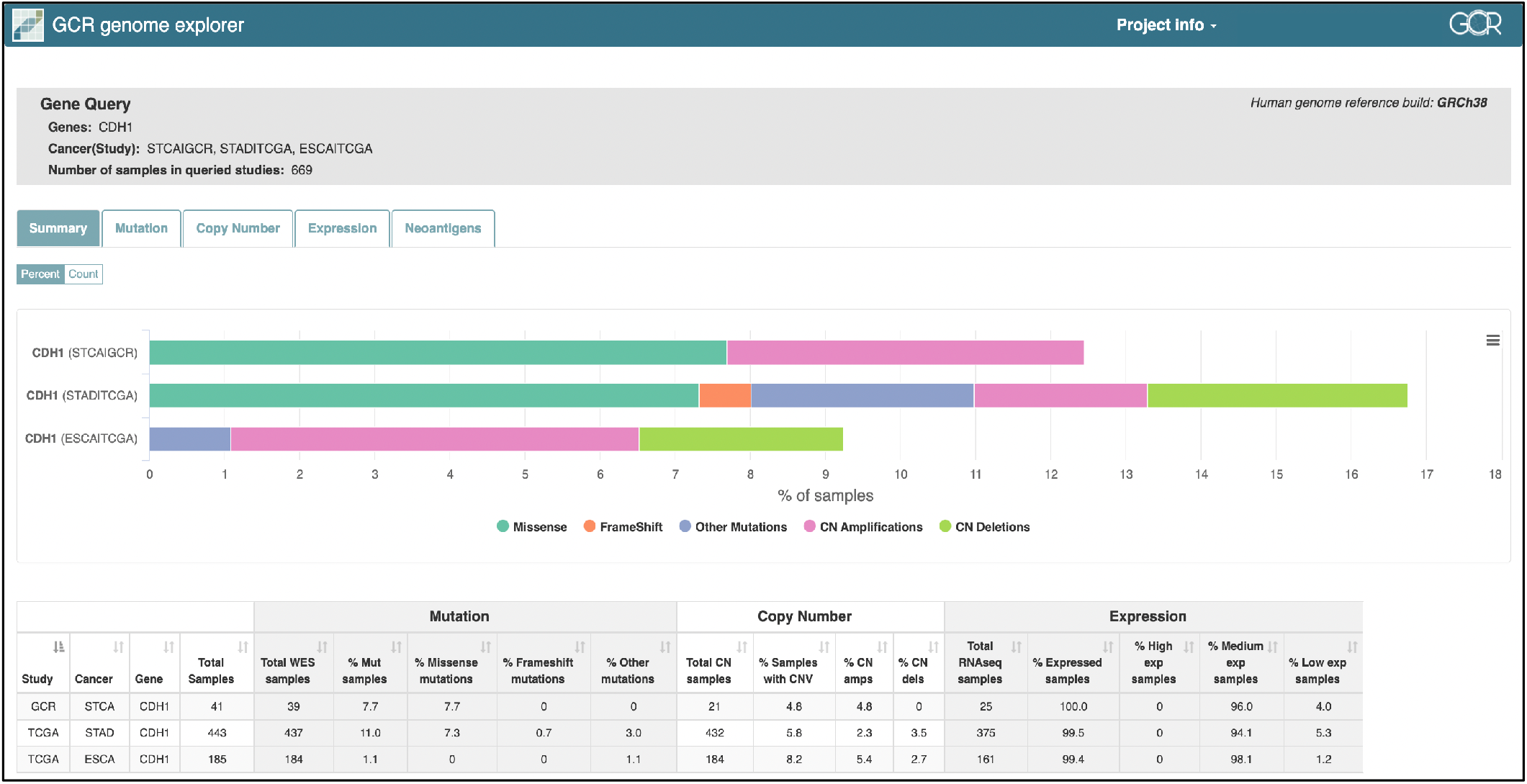
The GCR Genome-Explorer output of a “Gene” query across all study cohorts for CDH1. The “Summary” page displays counts and percentages of samples with CDH1 mutations, copy number variations, and expression levels. The “Mutation,” “Copy Number,” “Expression,” and “Neoantigen” tabs contain more detailed gene alterations among the studies’ patients.

The “Neoantigen” query gives the user the option to select an HLA allele type and view a list of the candidate neoantigens. For example, the most prevalent HLA-A type in the GCR-STCA study is A*02:01 (18.9%). Selecting this HLA type in the Neoantigen query produces a breakdown of all neoantigen candidates. They are described by their gene, chromosomal location, amino acid change and binding strength/rank based on their predicted properties in terms of MHC1 interaction **(Figure 3)**. Like the “Gene” query, the “Neoantigen” query function allows users to search data across multiple patient cohorts.

**Figure 3.**
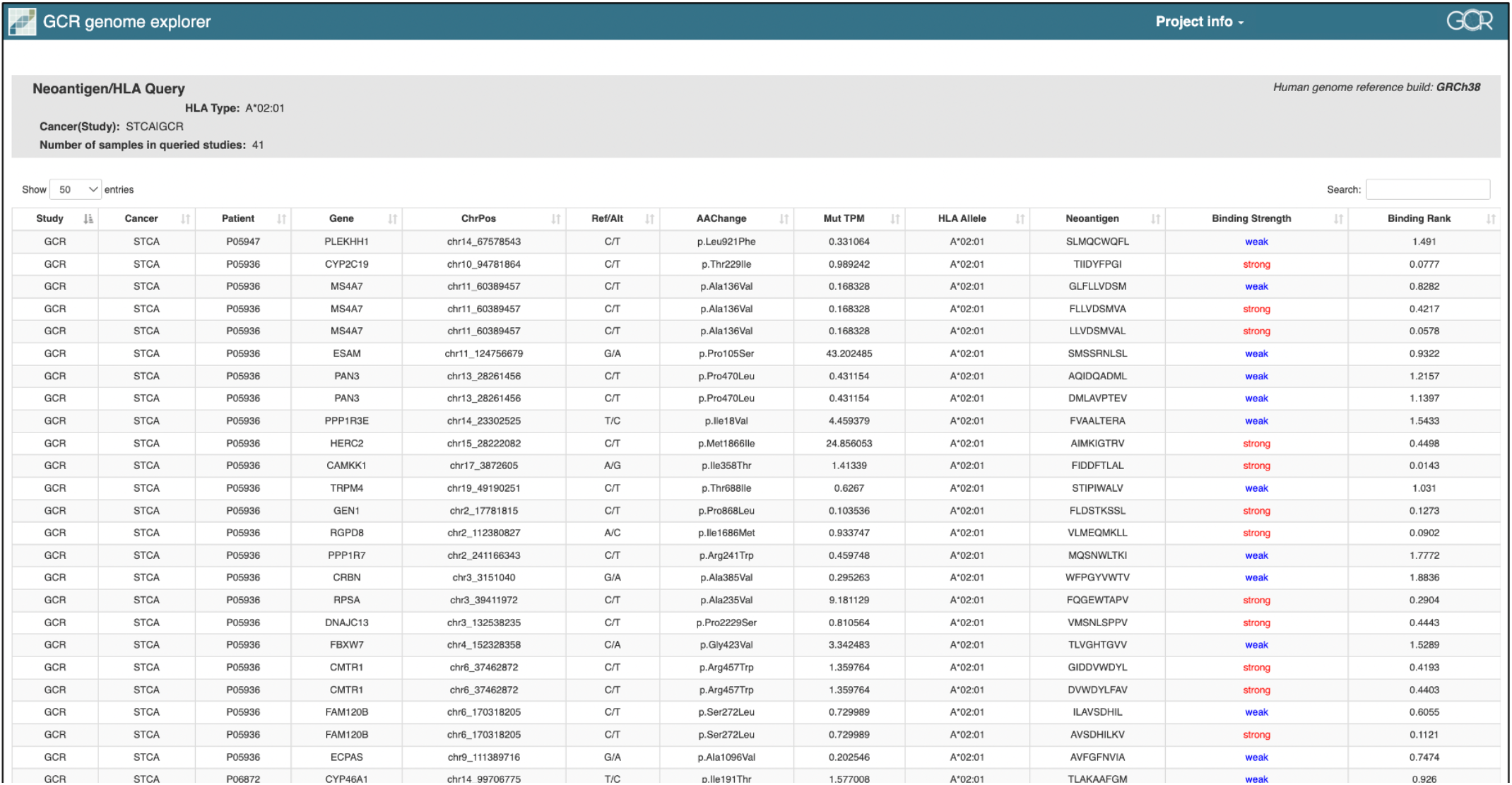
Example of the “Neoantigen” query for HLA type A*02:01 within the GCR-STCA cohort. The table describes specific gene mutations that lead to the production of a neoantigen and the predicted binding strength of the neoantigen to HLA allele A*02:01.

Lastly, the “Patient” query function provides users a way to see all available data for an individual patient in the database (**Figure 4**). The patient’s summary page includes a count of mutations and copy number variations, their HLA-A, -B, and -C types, and their clinical characteristics. Like the “Explore Study” feature and the “Gene” query, the “Patient” query arranges datasets into tabs. The “Mutation” tab details the patient’s unique mutations with chromosome position, reference sequence and alteration, variant type, and amino acid change. The “Copy Number” page lists genes with duplication and deletion events. In the “Gene Expression” tab, the user can view the expression level of a gene through FPKM and qualitative values of high, medium, and low. The gene list can be filtered to include only cancer-associated genes, oncogenes, or tumor suppressor genes. The next tab contains putative neoantigens described by position, amino acid change, and binding strength to a specific HLA allele. The final tab displays the patient’s microbiome, a taxonomy of microbial populations classified from kingdom to genus.

**Figure 4.**
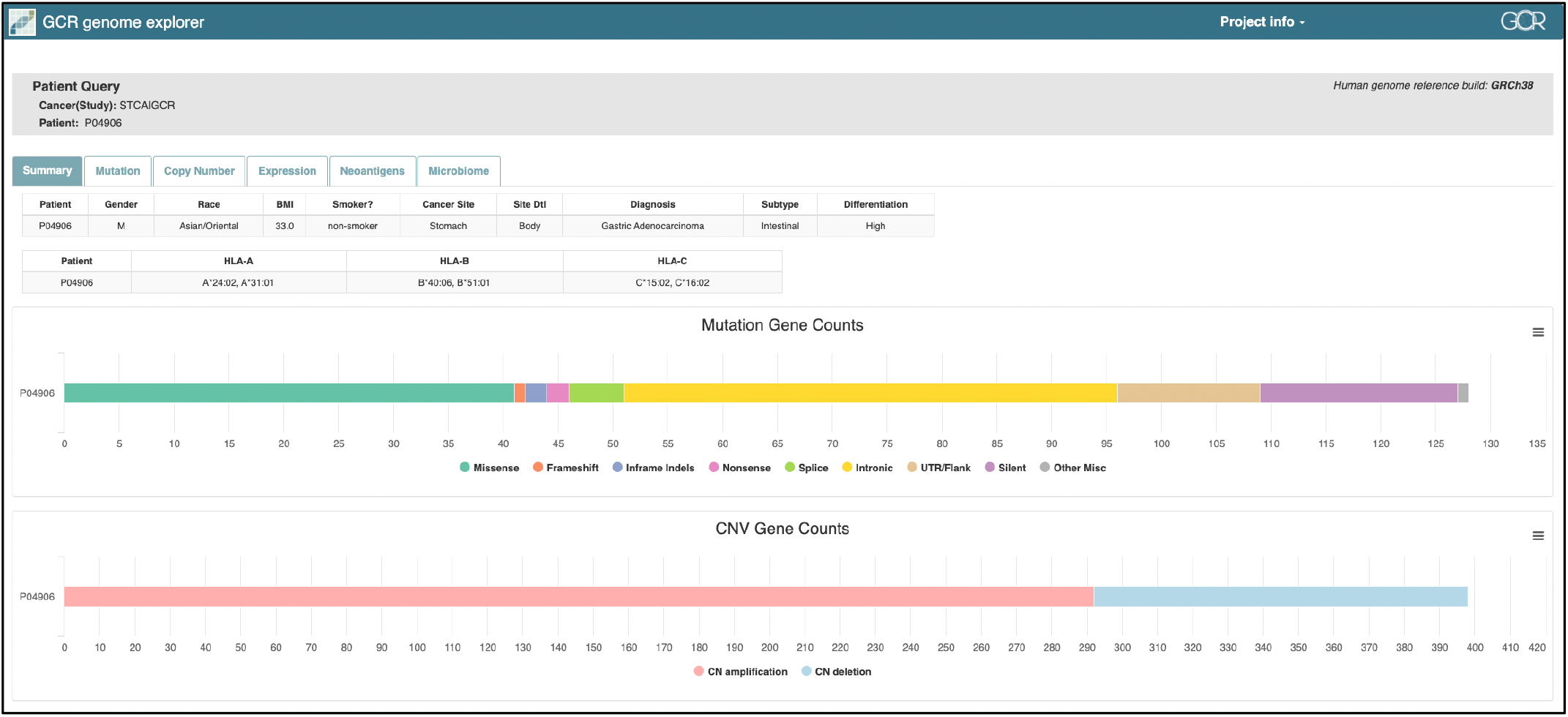
“Patient” query for P04906 from the GCR study. Multiple tabs present the entirety of clinical, genomic, and cellular data for this individual.

## DISCUSSION

The GCR’s goal is to catalyze research which will accelerate the prevention, detection, and treatment of this malignancy. Patients with GC, a family history of GC in a first-or second-degree relative, a germline *CDH1* mutation, or a combination of the three were invited to enroll. Recruitment efforts were facilitated by making enrollment simple and available to individuals who fulfill the eligibility criteria. Our national enrollment to patients outside of the northern California region has enabled us to increase the diversity of the participants. Through ongoing collaborations with other research groups and healthcare centers across the globe, we are continuing to build out a robust repository of clinical datasets, biospecimens and genomic data. The ongoing expansion of the GCR will provide an even more diverse cohort and broaden the utility of this data for research.

The GCR integrates epidemiology and genomic information from individuals at high risk for GC or patients diagnosed with this malignancy. GCR Genome Explorer users can conduct studies with this genomic information and take advantage of the sophisticated bioinformatic tools used to process it. For example, a researcher can pose a question, “I found that *H. pylori* infected patients often have downstream functional mutation in the gene *MUC1*; can this finding be replicated in another patient population?” The result is provided efficiently among the GCR population as well as other studies such as the TCGA. The information may be applicable to other cancer research studies as well.

While the pilot release of the Genome Explorer currently contains genomic data for 41 gastric tumors, our most recent enrollment will quadruple the number of samples available for genomic studies. This influx of new participants is providing additional clinical datasets such as pathology reports and other clinical metrics. On this expanded cohort we are conducting additional genomic studies which will greatly increase the overall number of tumors with genomic data. As the registry continues to accrue participants and tumor samples, we anticipate that there will be improved statistical power for future studies.

Since the inception of the GCR, other groups have compiled clinical or genomic data on GC patients. The Stomach Cancer Pooling **(StoP)** Project is an international consortium for epidemiological investigations into GC with 22 independent case-control studies participating.^18^ Organized in 2015, the StoP consortium conducted multiple studies to quantify the association between diet and lifestyle choices,^19-22^ socioeconomic status,^23^ and other factors^24-26^ with the risk of GC. In 2015, researchers identified discrete sources of epidemiological, clinicopathological, and molecular biological information from GC researchers and treatment centers in China.^27^ They standardized the data to produce the Database of Human Gastric Cancer.^27^ While valuable, neither of these cited resources address the current need for an easily accessible, comprehensive database that links detailed GC genomic analysis with clinical data. What differentiates the GCR from these other efforts comes from its integration of clinical and genomic data from GC patients, making it invaluable for future translational studies. Moreover, there is no single genomic database of GCs based on patients from the United States, a region with a lower prevalence of *H. pylori-*induced cancer.^28^

For this iteration of the GCR’s current registry we noted several limitations related to patient reporting. First, participants in the GCR completed varying degrees of their questionnaires. All participants who completed our online consent form were included in the final cohort. However, some participants did not complete every field of their questionnaire. Second, not all participants donated biological samples to the study. Some patients elected not to contribute. This issue was exacerbated by the COVID-19 pandemic during which many patients were unable to donate saliva samples given the biohazard issues. Third, using an online self-enrollment method limits participation to individuals with access to the internet. One study suggested that older individuals and those with lower self-rated internet ability are less likely to complete online enrollment processes.^29^ Finally, our ongoing recruitment is expanding the overall diversity of the cohort.

In summary, the GCR provides open access to a wealth of clinical and genomic information from a steadily expanding number of gastric tumors and individuals with hereditary predispositions for GC. We developed a framework for collating this information, analyzing tumor samples, and accessing this information via an online registry portal. Our results are openly accessible for independent discovery and validation studies.

## Data Availability

Primary data is available on the Gastric Genome Explorer website (https://gcregistry-explorer.stanford.edu/). Other data generated in this study are available within the article and its supplementary files.

## ABBREVIATIONS

GC: Gastric cancer
GCR: Gastric Cancer Registry
FFPE: Formalin fixed paraffin embedded
RNA-seq: RNA sequencing
HLA: Human leukocyte antigen
TCGA: The Cancer Genome Atlas
STAD: Stomach adenocarcinoma
ESCA: Esophageal carcinoma
StoP: Stomach Cancer Pooling Project

## ACKNOWLEDGEMENTS

The REDCap platform services at Stanford are subsidized by a) Stanford School of Medicine Research Office, and b) the National Center for Research Resources and the National Center for Advancing Translational Sciences, National Institutes of Health, through grant UL1 TR001085.

## DATA ACCESSIBILITY

Primary data is available on the Gastric Genome Explorer website (https://gcregistry-explorer.stanford.edu/). Other data generated in this study are available within the article and its supplementary data files.

## Notes

**FINANCIAL SUPPORT** The work was supported by the Gastric Cancer Foundation. Additional support to HPJ came from the Research Scholar Grant, RSG-13-297-01-TBG from the American Cancer Society, and the Clayville Foundation. RJH is supported by the National Cancer Institute of the National Institutes of Health under Award Number K08CA252635.

**CONFLICT OF INTEREST** The authors have no competing interests to declare.

### Competing Interest Statement

The authors have declared no competing interest.

### Funding Statement

The work was supported by the Gastric Cancer Foundation. Additional support to HPJ came from the Research Scholar Grant, RSG-13-297-01-TBG from the American Cancer Society, and the Clayville Foundation. RJH is supported by the National Cancer Institute of the National Institutes of Health under Award Number K08CA252635.

### Author Declarations

IRB of Stanford University gave ethical approval of this work (IRB-20285).

