## Supplemental File 1 for "The Gastric Cancer Registry: A Genomic Translational Resource for Multidisciplinary Research in Stomach Malignancies"

**SUPPLEMENTARY FILE 1**

**TITLE**

### The Gastric Cancer Registry: A Genomic Translational Resource for Multidisciplinary Research in Stomach Malignancies

**AUTHORS**

Alison F Almeda[1], Sue Grimes[1], HoJoon Lee[1], Stephanie U Greer[1], GiWon Shin[1], Madeline McNamara[1], Anna C Hooker[1], Maya Arce[1], Matthew Kubit[1], Marie Schauer[1], Paul Van Hummelen[1], Cindy Ma[1], Meredith Mills[1], Robert J Huang[2], Joo Ha Hwang[2], Manuel R Amieva[3], Summer Han[4], James M. Ford[1], Hanlee P. Ji[1*]

[1]Division of Oncology, Department of Medicine, Stanford University School of Medicine, Stanford, CA, 94305, United States

[2]Division of Gastroenterology and Hepatology, Department of Medicine, Stanford University School of Medicine, Stanford, CA, 94305, United States

[3]Division of Infectious Diseases, Department of Pediatrics, Stanford University School of Medicine, Stanford, CA, 94305, United States

[4]Department of Neurosurgery, Stanford University School of Medicine, Stanford, CA, 94305, United States

**CORRESPONDING AUTHOR**

Hanlee P. Ji

Division of Oncology, Department of Medicine – Stanford University School of Medicine

CCSR 2245, 269 Campus Drive

Stanford, CA 94305-5151

### Initial Gastric Cancer Registry Questionnaire

Welcome to the Gastric Cancer Registry!

We are collecting information from gastric cancer patients including:

- Lifestyle and family history (online)

- Blood, saliva and fixed tissue samples

Anyone who is 18 years or older with a personal or family history of gastric cancer and/or has tested positive for a CDH1 mutation may participate.

The consent form on the next page explains the study in more detail. After reading and agreeing to the consent, please complete this questionnaire as thoroughly as possible to participate in the registry.

Research study staff will contact you (please mark your preferred method of communication on the registration form) to arrange blood, saliva and tissue collection after you have completed the questionnaire. Please note that we are unable to return any individual results.

If you have any questions, please contact the Gastric Cancer Registry Team at or (650) 497-3619.

#### Registration Form

| | |
|---|---|
| Please check one of the following items: | ☐ I currently have or have had gastric cancer and am registering myself in the registry<br>☐ I have a family history of gastric cancer in a first or second degree relatives<br>☐ I have tested positive (pathogenic or deleterious) for a mutation in the CDH1 gene (also called E-cadherin)<br>☐ None of the above is true. This means you are not an eligible participant for the Gastric Cancer Registry and will end this survey please contact us with questions. |
| First degree relatives with gastric cancer: | ☐ Mother<br>☐ Father<br>☐ Brother<br>☐ Sister<br>☐ Son<br>☐ Daughter |
| Second degree MATERNAL (mother's side) relatives with gastric cancer: | ☐ Aunt<br>☐ Uncle<br>☐ Nephew<br>☐ Niece<br>☐ Grandfather<br>☐ Grandmother<br>☐ Half-sibling<br>☐ Grandchild |

| | |
|---|---|
| Second degree PATERNAL (father's side) relatives with gastric cancer: | ☐ Aunt<br>☐ Uncle<br>☐ Nephew<br>☐ Niece<br>☐ Grandfather<br>☐ Grandmother<br>☐ Half-sibling<br>☐ Grandchild |
| Your name: | __________________________________ |
| Mailing Address | |
| E-mail address<br>(only to be used by research staff to contact you) | __________________________________ |
| Phone Number (including area code) | __________________________________ |
| What is the best contact method from above (check one)? | ◯ Mailing Address<br>◯ E-mail Address<br>◯ Phone Number |
| How did you find the Gastric Cancer Registry? | ☐ Internet search<br>☐ Gastric Cancer Foundation Website<br>☐ My doctor<br>☐ Mercy General Hospital<br>☐ Gastric Cancer Support Group<br>☐ clinicaltrials.gov<br>☐ Other |
| At what institution/hospital does your doctor work? | __________________________________ |
| Please explain. If referred by a relative, please state their full name: | __________________________________________ |

#### Consent Form

CONSENT FORM

Protocol Director: Hanlee P. Ji, M.D.
Protocol Title: The Gastric Cancer Registry

Approval Date: February 10, 2021
Expiration Date: October 31, 2021

FOR QUESTIONS ABOUT THE STUDY, CONTACT: Dr. Hanlee Ji, 269 Campus Drive, CCSR 1120, Stanford, CA 94305, (650) 498-6000.

DESCRIPTION: You are invited to participate in this research study because you have been diagnosed with gastric cancer or have a relative who has had gastric cancer or you have a mutation in your CDH1 gene which increases your risk of gastric cancer.
Through this study we hope to learn more about patients with gastric cancer and those at an increased risk of gastric cancer, specifically unique patient characteristics and risk factors for developing gastric cancer. Your participation in this study gives us permission to collect clinical information from your medical record and store it in a database for future analysis with information about other patients with gastric cancer or those with a strong family history of gastric cancer. We will also collect other information from you regarding your lifestyle, family history and possible risk factors through a questionnaire. This study also requires that we collect a blood sample for future testing, done so during your regular medical care (i.e. when you have a regularly scheduled blood draw), and/or a saliva sample, collected easily at home. Your participation also allows us to request previously collected samples of gastric tumor and noncancerous stomach tissue from outside hospitals if not collected at Stanford. The tissue samples we request are from procedures done in the course of your treatment. Participation in this study will not require any additional tissue samples to be taken for research.
Your participation in this study is entirely voluntary. Your decision whether or not to participate will not prejudice you or your medical care. If you decide to participate, you are free to withdraw your consent, and to discontinue participation at any time without prejudice to you or effect on your medical care.

You will be asked to provide a sample of bodily fluids in the form of saliva and/or blood. Donating saliva is a non-invasive way to provide a sample that can be easily given at home. Our study staff will contact you to have a saliva sample collection kit sent to your mailing address. Additionally, we would like to collect a small blood sample (approximately 1.5 tablespoons). The blood will be taken with a needle from your arm at the same time you are having blood drawn for your standard clinic visit. If you live outside the Stanford area, our study staff will contact you to arrange your blood draw at a local facility and to have the blood shipped to Stanford.

You will also be asked to complete a detailed online questionnaire about your medical, social and family history to help us learn more about causes of gastric cancer. The questionnaire should take about 30 minutes to complete. A feedback survey will be sent automatically to you 1 month after you enroll in the study. You have the right to refuse any question.
In addition to collecting and examining blood samples, we will review your medical chart to obtain information about your medical history as it relates to your gastric cancer (if applicable), as well as request paraffin-embedded tumor and noncancerous stomach tissue from outside hospitals if not collected at Stanford previously.
RISKS AND BENEFITS: The risks associated with this study are slight discomfort or bruising from the blood draw. You will not benefit from the study, as this is not a treatment study. We cannot and do not guarantee or promise that you will receive any benefits from this study. Your decision whether or not to participate in this study will not affect your medical care.
TIME INVOLVEMENT: Your participation in this experiment will take approximately an hour and a half and 5 minutes to register for the study online; 15 minutes for the consent process, 20 minutes for saliva and blood collection, and 45 minutes to complete the patient questionnaires. Specimens will be banked for 50 years.

PAYMENTS: You will not be paid to participate in this study.

PARTICIPANT'S RIGHTS: If you have read this form and have decided to participate in this project, please understand your participation is voluntary and you have the right to withdraw your consent or discontinue participation at any time without penalty or loss of benefits to which you are otherwise entitled. You have the right to refuse to answer particular questions.

Your individual privacy will be maintained in all published and written data resulting from the study.

TISSUE SAMPLING FOR FUTURE RESEARCH
Research using tissues is an important way to try to understand human disease. You have been given this information because the investigators want to include your tissues in a research project and because they want to save the samples for future research. There are several things you should know before allowing your tissues to be studied.
Your tissues will be stored with a code number, not your medical record number and will be linked to the other data you have provided
You have the right to refuse to allow your tissues to be studied now or saved for future study. You may withdraw from this study at any time. The investigators might retain the identified samples, e.g., as part of your routine

clinical care, but not for additional research.

The results of the study of your samples will be used for research purposes only and you will not be told the results of the tests unless we discover something that would be of medical benefit for you or your family.

Any tissues you have donated which are used in research may result in new products, tests or discoveries. In some instances, these may have potential commercial value and may be developed and owned by the Investigators, Stanford University and/or others. However, donors of tissues do not retain any property rights to the materials. Therefore, you would not share in any financial benefits from these products, tests or discoveries.

TISSUE SAMPLING FOR GENETIC TESTING.

As part of the analysis on your samples, the investigators may do genetic testing. Genetic research is research that studies genes, including gene characteristics and gene versions that are transmitted by parents to children. Genetic research may include looking at information, such as personal appearance and biochemistry, gene sequences, genetic landmarks, individual and family medical histories, reactions to medications and responses to treatment. Genetic research raises certain questions about informing you of any results. Possible risks of knowing results include: anxiety; other psychological distress; and the possibility of insurance and job discrimination. A possible risk of not knowing includes being unaware of the need for treatment. These risks can change depending on the results of the research and whether there is a treatment or cure for a particular disease. The results of the study of your samples from this project will be used for research purposes only, and you will not be told the results of the tests.

Sometimes patients have been required to furnish information from genetic testing for health insurance, life insurance, and/or a job. A Federal law, the Genetic Information Nondiscrimination Act of 2008 (GINA), generally makes it illegal for health insurance companies, group health plans, and employers with 15 or more employees to discriminate against you based on your genetic information.

Information from analyses of your coded samples and your coded medical information will be put into public databases housed by Stanford University as well as one of the National Institutes of Health (NIH) databases along with information from the other research participants and will be used for future research. These databases will be accessible by the Internet. Only anonymous information from the analyses will be put in a completely public database, available to anyone on the Internet. Access to de-identified data will be granted according to the terms of the database and used for any research purpose.

No traditionally-used identifying information about you, such as your name, address, telephone number, or social security number, will be put into the public database. While the public database will not contain information that is traditionally used to identify you, people may develop ways in the future that would allow someone to link your genetic or medical information in our databases back to you. For example, someone could compare information in our databases with information from you (or a blood relative) in another database and be able to identify you (or your blood relative). It also is possible that there could be violations to the security of the computer systems used to store the codes linking your genetic and medical information to you.

However, your privacy is very important to us and we will use safety measures to protect it. Despite all of the safety measures that we will use, we cannot guarantee that your identity will never become known.

Authorization to Use Your Health Information
for Research Purposes

Because information about you and your health is personal and private, it generally cannot be used in this research study without your written authorization. If you agree to this form and continue with the following questionnaire, it will provide that authorization. The form is intended to inform you about how your health information will be used or disclosed in the study. Your information will only be used in accordance with this authorization form and the informed consent form and as required or allowed by law. Please read it carefully before agreeing with it.

What is the purpose of this research study and how will my health information be utilized in the study?
To study unique patient characteristics, risk factors, and outcomes of patients with gastric cancer.

Do I have to agree with this authorization form?
You do not have to agree with this authorization form. But if you do not, you will not be able to participate in this research study.

If I agree to this study, can I revoke it or withdraw from the research later?
If you decide to participate, you are free to withdraw your authorization regarding the use and disclosure of your health information (and to discontinue any other participation in the study) at any time. After any revocation, your health information will no longer be used or disclosed in the study, except to the extent that the law allows us to continue using your information (e.g., necessary to maintain integrity of research). If you wish to revoke your authorization for the research use or disclosure of your health information in this study, you must do so in writing. Please provide the written withdrawal to Dr. Hanlee Ji, 269 Campus Drive, CCSR 1120 Building, Stanford, CA.

What Personal Information Will Be Used or Disclosed?

Health information about you obtained from studying your blood sample, tissue sample, relevant information about you and your tumor obtained from the medical record, and information collected from the patient questionnaire.

Who May Use or Disclose the Information?
The following parties are authorized to use and/or disclose your health information in connection with this research study:

• The Protocol Director, Hanlee Ji, M.D.
• The Stanford University Administrative Panel on Human Subjects in Medical Research and any other unit of Stanford University as necessary
• Research Staff working on this study

Who May Receive or Use the Information?
The parties listed in the preceding paragraph may disclose your health information to the following persons and organizations for their use in connection with this research study:

• The Office for Human Research Protections in the U.S. Department of Health and Human Services

Your information may be re-disclosed by the recipients described above, if they are not required by law to protect the privacy of the information.

When will my authorization expire?
Your authorization for the use and/or disclosure of your health information will expire on February 22, 2111.

SPONSOR:

The V Foundation and the Gastric Cancer Foundation are providing financial support for this study.

CONTACT INFORMATION:
Questions, Concerns, or Complaints: If you have any questions, concerns or complaints about this research study, its procedures, risks and benefits, or alternative courses of treatment, you should ask the Protocol Director, Dr. Hanlee Ji. You may contact him now or later at (650) 498-6000.

Injury Notification: If you feel you have been hurt by being a part of this study, please contact the Protocol Director, Dr. Hanlee Ji at (650) 498-6000.

Independent Contact: If you are not satisfied with how this study is being conducted, or if you have any concerns, complaints, or general questions about the research or your rights as a participant, please contact the Stanford Institutional Review Board (IRB) to speak to someone independent of the research team at (650)-723-5244 or toll free at 1-866-680-2906. You can also write to the Stanford IRB, Stanford University, 1705 El Camino Real, Palo Alto, CA 94306.

EXPERIMENTAL SUBJECTS BILL OF RIGHTS: As a research participant you have the following rights. These rights include but are not limited to the participant's right to:
• be informed of the nature and purpose of the experiment;
• be given an explanation of the procedures to be followed in the medical experiment, and any drug or device to be utilized;
• be given a description of any attendant discomforts and risks reasonably to be expected;
• be given an explanation of any benefits to the subject reasonably to be expected, if applicable;
• be given a disclosure of any appropriate alternatives, drugs or devices that might be advantageous to the subject, their relative risks and benefits;
• be informed of the avenues of medical treatment, if any available to the subject after the experiment if complications should arise;
• be given an opportunity to ask questions concerning the experiment or the procedures involved;
• be instructed that consent to participate in the medical experiment may be withdrawn at any time and the subject may discontinue participation without prejudice;
• be given a copy of the dated consent form; and
• be given the opportunity to decide to consent or not to consent to a medical experiment without the intervention of any element of force, fraud, deceit, duress, coercion or undue influence on the subject's decision.

Please print a copy of this page for your records.

If you agree to participate in this research, please continue with the following questionnaire.

#### Demographics

Please complete this questionnaire as thoroughly as possible to participate in the registry. You can exit the survey and return to your place as needed, but remember to WRITE DOWN THE KEY CODE given upon exit because you will need that to get back into your survey.

Additionally, once you save a page and move to the next one, you cannot return to the previous page nor can you change your answers on previous pages, so PLEASE DOUBLE CHECK YOUR ANSWERS BEFORE YOU MOVE TO A NEW PAGE.

Many questions will ask for a date. You should select a date from the calendar drop-down. Note, you will have to select a specific day for it to work. If the exact day is unknown, please choose the first day of the known month and year.

If you have any questions, please contact the Research Assistant for the Gastric Cancer Registry, Alison Almeda at (650) 497-3619 or

1 Last name

__________________________________

timestamp

__________________________________

2 First name (use full given name rather than nicknames)

__________________________________

3 Middle initial

__________________________________

4 Date of Birth

__________________________________

5 Age

__________________________________

6 Sex

○ Female ○ Male

7 Country of birth (for United States, please write USA)

__________________________________

8 If country of birth is not United States, what year you did you permanently move to the USA?

__________________________________

9 Race (Select all that apply)

- ☐ White
- ☐ Black
- ☐ Native American/Alaska Native
- ☐ Asian
- ☐ Pacific Islander
- ☐ Other

Nationality/Country

__________________________________

10 Ethnicity

◯ Hispanic ◯ Non-hispanic

11 Are you of Ashkenazi Jewish decent?

◯ Yes ◯ No

12 Height (in inches)

__________________________________

13 Weight (in pounds)

__________________________________

14 If you have had a gastrectomy (part or all of your stomach removed), what was your weight prior to your surgery (in pounds)?

__________________________________

#### Social History

15 Please choose the highest level of education you have completed.

◯ Elementary School ◯ High School ◯ College ◯ Graduate School

16 Current Marital Status

◯ Single ◯ Married/Partnered ◯ Divorced ◯ Separated ◯ Widowed/Widower

17 Current Employment Status

◯ Employed Full-time
◯ Employed Part-time
◯ Unemployed
◯ Retired
◯ Disability
◯ Working at Home
◯ Student

18 Total annual income of your household:

◯ $0-$25,000
◯ $25,001-$50,000
◯ $50,001-$75,000
◯ $75,001-$100,000
◯ $100,000 and above

19 Number of persons aged 18 or older who are currently living in your household.

________________________________

20 Which of the following best characterizes the area around your home? If you live in two homes, select the answer for the home in which you spend the most time.

◯ Urban ◯ Suburban ◯ Rural

#### Tobacco History

21 Have you ever smoked cigarettes?
◯ Yes
◯ No

21a How old were you when you started smoking cigarettes?

__________________________________

21b During the time that you smoked, what is the number of cigarettes per week you smoked?

__________________________________

21c Do you currently smoke cigarettes (within the last month)?
◯ Yes
◯ No

21d What was the date that you stopped smoking cigarettes (best approximation)?

__________________________________

22 Have you ever smoked a pipe?
◯ Yes
◯ No

22a How old were you when you started smoking a pipe?
__________________________________

22b During the time that you smoked, what is the average number of ounces of tobacco per week you smoked?
__________________________________

22c Do you currently smoke a pipe (within the last month)?
◯ Yes
◯ No

22d What was the date you stopped (best approximation)?
__________________________________

23 Have you ever smoked cigars?
◯ Yes
◯ No

23a How old were you when you started smoking cigars?
__________________________________

23b During the time that you smoked, what is the number of cigars per month that you smoked?
__________________________________

23c Do you currently smoke cigars (within the last month)?
◯ Yes
◯ No

23d What was the date that you stopped (best approximation)?
__________________________________

#### Alcohol History

24 Have you ever or do you currently drink alcohol?

- ◯ Yes, but only in the past
- ◯ Yes, currently
- ◯ No, never

25 How many alcoholic beverages (beer, wine, mixed drinks, etc) do you CURRENTLY consume?

- ◯ None
- ◯ Less than 1 per week
- ◯ 1-2 drinks per week
- ◯ 3-4 drinks per week
- ◯ 5-8 drinks per week
- ◯ 9-12 drinks per week
- ◯ 13-15 drinks per week
- ◯ More than 15 drinks per week

26 If your alcohol intake in the past was different from now, how many alcoholic beverages (beer, wine, mixed drinks, etc) did you consume IN THE PAST?

- ◯ None
- ◯ Less than 1 per week
- ◯ 1-2 drinks per week
- ◯ 3-4 drinks per week
- ◯ 5-8 drinks per week
- ◯ 9-12 drinks per week
- ◯ 13-15 drinks per week
- ◯ More than 15 drinks per week

#### Medications

| | | |
|---|---|---|
| 27 | Have you previously or do you currently use aspirin regularly? | ◯ Yes<br>◯ No |
| 27a | What dosage (in mg)? | __________________ |
| 28 | Have you previously or do you currently use non-steroidal anti-inflammatory drugs (Ibuprofen, Advil, Motrin, Aleve, etc) regularly? | ◯ Yes<br>◯ No |
| 29 | Have you previously or do you currently use blood thinners (Coumadin, Lovenox, etc) regularly? | ◯ Yes<br>◯ No |
| 30 | Have you previously or do you currently use an antacid (Maalox, Rolaids, Tums, etc) regularly? | ◯ Yes<br>◯ No |
| 31 | Have you previously or do you currently use an H2 blocker (Ranitidine, Zantac, Cimetidine, Tagamet, Famotidine, Pepcid, etc) regularly? | ◯ Yes<br>◯ No |
| 32 | Have you previously or do you currently use a Proton Pump Inhibitor (Omeprazole, Prilosec, Lansoprazole, Prevacid, Nexium, etc) regularly? | ◯ Yes<br>◯ No |
| 33 | Have you previously or do you currently use a multivitamin regularly? | ◯ Yes<br>◯ No |

34 Please list any other medications, vitamins, supplements, and other mineral products you're currently taking:

#### Medical History

35 Do you have a history of diabetes or high blood sugar?
◯ No
◯ Yes
◯ Unknown

35a What year were you first diagnosed with diabetes or high blood sugar?
__________________________________

35b For child-bearing women, was this diagnosed as gestational diabetes?
◯ No
◯ Yes
◯ Unknown

36 Do you have a history of high cholesterol?
◯ No
◯ Yes
◯ Unknown

36a What year were you first diagnosed with high cholesterol?
__________________________________

37 Do you have a history of high triglycerides?
◯ No
◯ Yes
◯ Unknown

37a What year were you first diagnosed with high triglycerides?
__________________________________

38 Do you have a history of high blood pressure?
◯ No
◯ Yes
◯ Unknown

38a What year were you first diagnosed with high blood pressure?
__________________________________

39 In the past have you ever had any of the following types cancer (select ALL that apply).

☐ Anus
☐ Appendix
☐ Bile Ducts
☐ Bladder
☐ Brain
☐ Breast
☐ Cervical
☐ Colon (Large Intestine)
☐ Colorectal
☐ Esophagus
☐ Gallbladder
☐ Hodgkin's Lymphoma
☐ Kidney
☐ Leukemia
☐ Liver
☐ Lung
☐ Melanoma
☐ Mouth/throat
☐ Non-Hodgkin's Lymphoma
☐ Ovary
☐ Pancreas
☐ Pancreatic Islet Cell
☐ Parathyroid
☐ Pheochromocytoma
☐ Pituitary
☐ Prostate
☐ Rectum
☐ Small intestine
☐ Stomach
☐ Thyroid
☐ Uterine/Endometrial
☐ Unknown
☐ Other

39a Please specify (other): ____________________

39b Please specify which year (and for which diagnosis if more than one) you were diagnosed with the cancer from above.

40 At what age did you have your first period? ____________________

41 Are you still menstruating?

◯ Yes
◯ No

41a At what age did you stop menstruating (best approximation)? ____________________

41b Why did your period stop?

◯ Natural menopause
◯ Surgical menopause (had surgery to remove your ovaries and/or uterus)
◯ Chemotherapy-induced menopause
◯ Currently pregnant or nursing

42 Have you ever been pregnant?

◯ Yes
◯ No

42a How many times have you been pregnant?

________________________________

42b At what age did you have your first full-term pregnancy?

________________________________

43 Have you ever used birth control pills or other hormonal contraceptives?

- ◯ Yes
- ◯ No

43a How many months, in total, have you used birth control pills or other hormonal contraceptives?

________________________________

44 Have you ever used hormone replacement therapy (HRT)?

- ◯ Yes
- ◯ No

44a For how many months, in total, have you used HRT?

________________________________

44b What type of HRT did you use (check all that apply)?

- ☐ Estrogen alone
- ☐ Progesterone alone
- ☐ Both estrogen and progesterone
- ☐ Unknown

#### Gastric Cancer Related Risk Factors

45 Have you ever been diagnosed as having Epstein-Barr virus?
- ◯ No
- ◯ Yes
- ◯ Don't Know

45a What year was the diagnosis made?

__________________________

46 Have you ever been tested for Helicobacter pylori (H. pylori)?
- ◯ No
- ◯ Yes
- ◯ Don't Know

46b In what year did the test occur?

__________________________

46a Was the test positive?
- ◯ No
- ◯ Yes
- ◯ Don't Know

47 What is your blood type?
- ◯ A
- ◯ B
- ◯ AB
- ◯ O
- ◯ I don't know my blood type

48 Have you ever had gastric surgery for reasons other than gastric cancer?
- ◯ No
- ◯ Yes
- ◯ Don't Know

48a What year did you have gastric surgery (unrelated to your gastric cancer diagnosis)?

__________________________

49 Have you ever been diagnosed with a gastric ulcer?
- ◯ No
- ◯ Yes
- ◯ Don't Know

49a What year was the diagnosis?

__________________________

50 Have you ever been diagnosed with gastric polyps?
- ◯ No
- ◯ Yes
- ◯ Don't Know

50a What year was the diagnosis?

__________________________

51 Have you ever been diagnosed with gastroesophageal reflux disease (GERD)?
- ◯ No
- ◯ Yes
- ◯ Don't Know

51a What year was the diagnosis?

__________________________

52 Have you ever been diagnosed with pernicious anemia (your body has trouble absorbing vitamin B12)?
- ◯ No
- ◯ Yes
- ◯ Don't Know

52a What year was the diagnosis?

__________________________________

53 Have you ever been diagnosed with Menetrier's disease (folds of extra tissue in the stomach wall)?

- ◯ No
- ◯ Yes
- ◯ Don't Know

53a What year was the diagnosis?

__________________________________

54 Have you ever had an endoscopy?

- ◯ No
- ◯ Yes, just one
- ◯ Yes, more than one
- ◯ Don't Know

54a What was the date of your last endoscopy?

__________________________________

54b What were the findings of your last endoscopy?

- ☐ Normal
- ☐ Cancer
- ☐ Polyps
- ☐ Other

What type of cancer?

- ◯ Anus
- ◯ Appendix
- ◯ Bile Ducts
- ◯ Bladder
- ◯ Brain
- ◯ Breast
- ◯ Cervical
- ◯ Colon (Large Intestine)
- ◯ Colorectal
- ◯ Esophagus
- ◯ Gallbladder
- ◯ Hodgkin's Lymphoma
- ◯ Kidney
- ◯ Leukemia
- ◯ Liver
- ◯ Lung
- ◯ Melanoma
- ◯ Mouth/throat
- ◯ Non-Hodgkin's Lymphoma
- ◯ Ovary
- ◯ Pancreas
- ◯ Pancreatic Islet Cell
- ◯ Parathyroid
- ◯ Pheochromocytoma
- ◯ Pituitary
- ◯ Prostate
- ◯ Rectum
- ◯ Small intestine
- ◯ Stomach
- ◯ Thyroid
- ◯ Uterine/Endometrial
- ◯ Unknown
- ◯ Other

How many polyps were found?

__________________________________

Please describe:

54c Please list the rest of your endoscopies. Include date, result (normal, polyps, cancer, other) and describe the result in more detail if known. For example: 05/15/2007, normal; 05/02/2006, gastric cancer

55 Have you ever had a colonoscopy?

- ◯ No
- ◯ Yes, just one
- ◯ Yes, more than one
- ◯ Don't Know

55a What was the date of your last colonoscopy?

__________________________________

55b What were the findings of your last colonoscopy?

- ☐ Normal
- ☐ Cancer
- ☐ Polyps
- ☐ Other

What type of cancer?

- ◯ Anus
- ◯ Appendix
- ◯ Bile Ducts
- ◯ Bladder
- ◯ Brain
- ◯ Breast
- ◯ Cervical
- ◯ Colon (Large Intestine)
- ◯ Colorectal
- ◯ Esophagus
- ◯ Gallbladder
- ◯ Hodgkin's Lymphoma
- ◯ Kidney
- ◯ Leukemia
- ◯ Liver
- ◯ Lung
- ◯ Melanoma
- ◯ Mouth/throat
- ◯ Non-Hodgkin's Lymphoma
- ◯ Ovary
- ◯ Pancreas
- ◯ Pancreatic Islet Cell
- ◯ Parathyroid
- ◯ Pheochromocytoma
- ◯ Pituitary
- ◯ Prostate
- ◯ Rectum
- ◯ Small intestine
- ◯ Stomach
- ◯ Thyroid
- ◯ Uterine/Endometrial
- ◯ Unknown
- ◯ Other

---

How many polyps were found?

__________________________________

---

Please describe:

---

55c Please list the rest of your colonoscopies.  Include date, result (normal, polyps, cancer, other) and describe the result in more detail if known. For example: 11/01/2005, normal; 10/28/2000, normal; 09/01/1995, 3 polyps

---

56 Have you ever genetic testing for hereditary cancer syndromes?

- ◯ Yes
- ◯ No

| | | |
|---|---|---|
| 56a | What type of testing have you had done? | ☐ BRCA1 and BRCA2 (Breast/Ovarian Cancer)<br>☐ HNPCC (Hereditary Non-Polyposis Colon Cancer Disorder). Genes include MSH2, MLH1, MSH6, PMS2.<br>☐ APC (Familial adenomatous polyposis)<br>☐ MYH also known as MUTYH (Familial adenomatous polyposis)<br>☐ PTEN (Cowden Syndrome)<br>☐ P53 (Li-Fraumeni Syndrome)<br>☐ CDH1 also known as E-Cadherin (Hereditary Gastric Cancer)<br>☐ Other |
| | Please specify gene, result of testing (positive for a deleterious mutation, negative, variant of uncertain or unknown significance, or unknown), and specific mutation site if found. | |
| | What was the result of the BRCA1 and BRCA2 testing? | ◯ Positive for a deleterious mutation<br>◯ Negative<br>◯ Variant of unknown significance or inconclusive<br>◯ I don't know |
| | Please enter gene and specific site of mutation (if known): | __________________________ |
| | What was the result of the HNPCC gene testing (would have been some combination of one or more of the following genes: MSH2, MLH1, MSH6, PMS2)? | ◯ Positive for a deleterious mutation<br>◯ Negative<br>◯ Variant of unknown significance or inconclusive<br>◯ I don't know |
| | Please enter gene and specific site of mutation (if known): | __________________________ |
| | What was the result of the APC testing? | ◯ Positive for a deleterious mutation<br>◯ Negative<br>◯ Variant of unknown significance or inconclusive<br>◯ I don't know |
| | Please enter specific site (if known): | __________________________ |
| | What was the result of the MYH (or MUTYH) testing? | ◯ Positive for a deleterious mutation<br>◯ Negative<br>◯ Variant of unknown significance or inconclusive<br>◯ I don't know |
| | Please enter specific site (if known): | __________________________ |
| | What was the PTEN testing result? | ◯ Positive for a deleterious mutation<br>◯ Negative<br>◯ Variant of unknown significance or inconclusive<br>◯ I don't know |
| | Please enter specific site (if known): | __________________________ |

| | |
|---|---|
| What was the result of the P53 testing? | ◯ Positive for a deleterious mutation<br>◯ Negative<br>◯ Variant of unknown significance or inconclusive<br>◯ I don't know |
| Please enter specific site (if known): | __________________________________ |
| What was the result of the CDH1 (or E-Cadherin) testing? | ◯ Positive for a deleterious mutation<br>◯ Negative<br>◯ Variant of unknown significance or inconclusive<br>◯ I don't know |
| Please enter specific site (if known): | __________________________________ |

57 Are there any other risk factors you think we should have asked about?

#### Gastric Cancer History

| | | |
|---|---|---|
| | Have you ever been diagnosed with gastric cancer or CDH1 mutation? | ◯ Yes<br>◯ No |
| 58 | When did you first go to any doctor with symptoms related to your gastric cancer (approximate as closely as possible)? | __________________ |
| 59 | What were these symptoms and when did they start?<br>For example: (heartburn: 04/15/2004, stomach pain: 05/01/2004, etc). Approximate as best you can on the dates; if only month and year are known, use the first day of the month. | |
| 60 | What is the approximate date of your gastric cancer diagnosis? | __________________ |
| 61 | What was the location of your tumor? | ◯ Stomach<br>◯ Gastroesophageal (GE) junction<br>◯ I don't know |
| 62 | Have you had surgery for your gastric cancer? | ◯ Yes<br>◯ No |
| 62a | What is the approximate date of your surgery? | __________________ |
| 62b | Where did you have surgery? Please include the name of the hospital and the city and state it is located in. | |
| 63 | Have you had any radiation therapy for your gastric cancer? | ◯ Yes<br>◯ No |
| 63a | Where did you have your radiation therapy? Please include the hospital or doctor's office and the city and state they were located in. List all locations if applicable. | |
| 64 | Have you had any chemotherapy for your gastric cancer? | ◯ Yes<br>◯ No |
| 64a | Where did you have your chemotherapy? Please include the name of the hospital and/or doctor's office and the city and state they are located in. | |

| | | |
|---|---|---|
| | Was/is your gastric cancer HER2 positive (this would mean you may have taken a drug called Herceptin)? | ◯ No<br>◯ Yes<br>◯ Unknown |
| | Have you ever taken or are you currently taking Herceptin? | ◯ No<br>◯ Yes<br>◯ Unknown |
| 65 | Have you had any alternative treatments (acupuncture, intuitive healing, healing touch, etc.)? | ◯ Yes<br>◯ No |

65a Please describe the types of alternative treatments you have had:

#### Family History

| | | |
|---|---|---|
| 66 | Are you adopted? | ◯ Yes<br>◯ No |
| 66a | Do you know about the medical history of your biological relatives? | ◯ Yes<br>◯ No. Please describe only your children below, then save and continue to next page. |
| 67 | How many full siblings do you have (full siblings share both biologic parents)? | __________________________ |
| 68 | How many children do you have? | __________________________ |
| 69 | Is your mother still alive? | ◯ Yes<br>◯ No<br>◯ Unknown |
| 69a | What is your mother's current age or at what age did she die? | __________________________ |
| 69b | Has your mother ever had cancer? | ◯ Yes<br>◯ No<br>◯ Unknown |

69c What type of cancer? If more than one, include most recent here, and see below. If other, please describe below.

- ◯ Anus
- ◯ Appendix
- ◯ Bile Ducts
- ◯ Bladder
- ◯ Brain
- ◯ Breast
- ◯ Cervical
- ◯ Colon (Large Intestine)
- ◯ Colorectal
- ◯ Esophagus
- ◯ Gallbladder
- ◯ Hodgkin's Lymphoma
- ◯ Kidney
- ◯ Leukemia
- ◯ Liver
- ◯ Lung
- ◯ Melanoma
- ◯ Mouth/throat
- ◯ Non-Hodgkin's Lymphoma
- ◯ Ovary
- ◯ Pancreas
- ◯ Pancreatic Islet Cell
- ◯ Parathyroid
- ◯ Pheochromocytoma
- ◯ Pituitary
- ◯ Prostate
- ◯ Rectum
- ◯ Small intestine
- ◯ Stomach
- ◯ Thyroid
- ◯ Uterine/Endometrial
- ◯ Unknown
- ◯ Other

69d What was her age at diagnosis?

______________________________

69e If more than one cancer, please include any additional cancer types and ages at diagnoses below. Please also elaborate if "other" was selected above as cancer type.

70 Is your father still alive?

- ◯ Yes
- ◯ No
- ◯ Unknown

70a What is your father's current age or at what age did he die?

______________________________

70b Has your father ever had cancer?

- ◯ Yes
- ◯ No
- ◯ Unknown

70c What type of cancer? If more than one, include most recent cancer here, and see below. If other, please describe below.

- ◯ Anus
- ◯ Appendix
- ◯ Bile Ducts
- ◯ Bladder
- ◯ Brain
- ◯ Breast
- ◯ Cervical
- ◯ Colon (Large Intestine)
- ◯ Colorectal
- ◯ Esophagus
- ◯ Gallbladder
- ◯ Hodgkin's Lymphoma
- ◯ Kidney
- ◯ Leukemia
- ◯ Liver
- ◯ Lung
- ◯ Melanoma
- ◯ Mouth/throat
- ◯ Non-Hodgkin's Lymphoma
- ◯ Ovary
- ◯ Pancreas
- ◯ Pancreatic Islet Cell
- ◯ Parathyroid
- ◯ Pheochromocytoma
- ◯ Pituitary
- ◯ Prostate
- ◯ Rectum
- ◯ Small intestine
- ◯ Stomach
- ◯ Thyroid
- ◯ Uterine/Endometrial
- ◯ Unknown
- ◯ Other

70d What was his age at diagnosis?

__________________________________

70e If more than one cancer, please include any additional cancer types and ages at diagnoses below. Please also elaborate if "other" was selected above as cancer type.

Now we would like to ask specific questions about your children and siblings including their age and whether or not they have ever had cancer. If you do not know their exact age, please approximate as best you can.

Please complete the following for ALL OF YOUR FULL SIBLINGS AND CHILDREN REGARDLESS OF THEIR CANCER HISTORY.

| | | |
|---|---|---|
| 71 | Please select a first-degree relative (please only describe FULL siblings, meaning siblings who share both biological parents with you): | ◯ Sibling 1<br>◯ Sibling 2<br>◯ Sibling 3<br>◯ Sibling 4<br>◯ Sibling 5<br>◯ Sibling 6<br>◯ Sibling 7<br>◯ Sibling 8<br>◯ Sibling 9<br>◯ Sibling 10<br>◯ Child 1<br>◯ Child 2<br>◯ Child 3<br>◯ Child 4<br>◯ Child 5<br>◯ Child 6<br>◯ Child 7<br>◯ Child 8<br>◯ Child 9<br>◯ Child 10<br>◯ I do not have anymore first degree relatives to describe |
| | Sex: | ◯ Male<br>◯ Female |
| | Is he/she still alive? | ◯ Yes<br>◯ No<br>◯ Unknown |
| | What is his/her current age or at what age did he/she die? | __________________________________ |
| | Has he/she ever had cancer? | ◯ Yes<br>◯ No<br>◯ Unknown |

| | |
|---|---|
| What type of cancer did he/she have? If more than one, include most recent cancer here, and see below. If other, please describe below. | ◯ Anus<br>◯ Appendix<br>◯ Bile Ducts<br>◯ Bladder<br>◯ Brain<br>◯ Breast<br>◯ Cervical<br>◯ Colon (Large Intestine)<br>◯ Colorectal<br>◯ Esophagus<br>◯ Gallbladder<br>◯ Hodgkin's Lymphoma<br>◯ Kidney<br>◯ Leukemia<br>◯ Liver<br>◯ Lung<br>◯ Melanoma<br>◯ Mouth/throat<br>◯ Non-Hodgkin's Lymphoma<br>◯ Ovary<br>◯ Pancreas<br>◯ Pancreatic Islet Cell<br>◯ Parathyroid<br>◯ Pheochromocytoma<br>◯ Pituitary<br>◯ Prostate<br>◯ Rectum<br>◯ Small intestine<br>◯ Stomach<br>◯ Thyroid<br>◯ Uterine/Endometrial<br>◯ Unknown<br>◯ Other |
| What was his/her age at diagnosis? | ______________________________ |

If more than one cancer, please include any additional cancer types and ages of diagnoses below. Please also elaborate if "other" was selected above as cancer type.

| | |
|---|---|
| Please select a different first-degree relative (please only describe FULL siblings, meaning siblings who share both biological parents with you): | ◯ Sibling 1<br>◯ Sibling 2<br>◯ Sibling 3<br>◯ Sibling 4<br>◯ Sibling 5<br>◯ Sibling 6<br>◯ Sibling 7<br>◯ Sibling 8<br>◯ Sibling 9<br>◯ Sibling 10<br>◯ Child 1<br>◯ Child 2<br>◯ Child 3<br>◯ Child 4<br>◯ Child 5<br>◯ Child 6<br>◯ Child 7<br>◯ Child 8<br>◯ Child 9<br>◯ Child 10<br>◯ I do not have anymore first degree relatives to describe |
| Sex: | ◯ Male<br>◯ Female |
| Is he/she still alive? | ◯ Yes<br>◯ No<br>◯ Unknown |
| What is his/her current age or at what age did he/she die? | __________________________________ |
| Has he/she ever had cancer? | ◯ Yes<br>◯ No<br>◯ Unknown |

| | |
|---|---|
| What type of cancer did he/she have? If more than one, include most recent cancer here, and see below. If other, please describe below. | ◯ Anus<br>◯ Appendix<br>◯ Bile Ducts<br>◯ Bladder<br>◯ Brain<br>◯ Breast<br>◯ Cervical<br>◯ Colon (Large Intestine)<br>◯ Colorectal<br>◯ Esophagus<br>◯ Gallbladder<br>◯ Hodgkin's Lymphoma<br>◯ Kidney<br>◯ Leukemia<br>◯ Liver<br>◯ Lung<br>◯ Melanoma<br>◯ Mouth/throat<br>◯ Non-Hodgkin's Lymphoma<br>◯ Ovary<br>◯ Pancreas<br>◯ Pancreatic Islet Cell<br>◯ Parathyroid<br>◯ Pheochromocytoma<br>◯ Pituitary<br>◯ Prostate<br>◯ Rectum<br>◯ Small intestine<br>◯ Stomach<br>◯ Thyroid<br>◯ Uterine/Endometrial<br>◯ Unknown<br>◯ Other |
| What was his/her age at diagnosis? | ______________________________ |

If more than one cancer, please include any additional cancer types and ages of diagnoses below. Please also elaborate if "other" was selected above as cancer type.

| | |
|---|---|
| Please select a different first-degree relative (please only describe FULL siblings, meaning siblings who share both biological parents with you): | ◯ Sibling 1<br>◯ Sibling 2<br>◯ Sibling 3<br>◯ Sibling 4<br>◯ Sibling 5<br>◯ Sibling 6<br>◯ Sibling 7<br>◯ Sibling 8<br>◯ Sibling 9<br>◯ Sibling 10<br>◯ Child 1<br>◯ Child 2<br>◯ Child 3<br>◯ Child 4<br>◯ Child 5<br>◯ Child 6<br>◯ Child 7<br>◯ Child 8<br>◯ Child 9<br>◯ Child 10<br>◯ I do not have anymore first degree relatives to describe |
| Sex: | ◯ Male<br>◯ Female |
| Is he/she still alive? | ◯ Yes<br>◯ No<br>◯ Unknown |
| What is his/her current age or at what age did he/she die? | __________________________ |
| Has he/she ever had cancer? | ◯ Yes<br>◯ No<br>◯ Unknown |

What type of cancer did he/she have? If more than one, include most recent cancer here, and see below. If other, please describe below.

- ◯ Anus
- ◯ Appendix
- ◯ Bile Ducts
- ◯ Bladder
- ◯ Brain
- ◯ Breast
- ◯ Cervical
- ◯ Colon (Large Intestine)
- ◯ Colorectal
- ◯ Esophagus
- ◯ Gallbladder
- ◯ Hodgkin's Lymphoma
- ◯ Kidney
- ◯ Leukemia
- ◯ Liver
- ◯ Lung
- ◯ Melanoma
- ◯ Mouth/throat
- ◯ Non-Hodgkin's Lymphoma
- ◯ Ovary
- ◯ Pancreas
- ◯ Pancreatic Islet Cell
- ◯ Parathyroid
- ◯ Pheochromocytoma
- ◯ Pituitary
- ◯ Prostate
- ◯ Rectum
- ◯ Small intestine
- ◯ Stomach
- ◯ Thyroid
- ◯ Uterine/Endometrial
- ◯ Unknown
- ◯ Other

---

What was his/her age at diagnosis?

__________________________________

---

If more than one cancer, please include any additional cancer types and ages of diagnoses below. Please also elaborate is "other" was selected above as cancer type.

| | |
|---|---|
| Please select a different first-degree relative (please only describe FULL siblings, meaning siblings who share both biological parents with you): | ◯ Sibling 1<br>◯ Sibling 2<br>◯ Sibling 3<br>◯ Sibling 4<br>◯ Sibling 5<br>◯ Sibling 6<br>◯ Sibling 7<br>◯ Sibling 8<br>◯ Sibling 9<br>◯ Sibling 10<br>◯ Child 1<br>◯ Child 2<br>◯ Child 3<br>◯ Child 4<br>◯ Child 5<br>◯ Child 6<br>◯ Child 7<br>◯ Child 8<br>◯ Child 9<br>◯ Child 10<br>◯ I do not have anymore first degree relatives to describe |
| Sex: | ◯ Male<br>◯ Female |
| Is he/she still alive? | ◯ Yes<br>◯ No<br>◯ Unknown |
| What is his/her current age or at what age did he/she die? | ______________________________ |
| Has he/she ever had cancer? | ◯ Yes<br>◯ No<br>◯ Unknown |

| | |
|---|---|
| What type of cancer did he/she have? If more than one, include most recent cancer here, and see below. If other, please describe below. | ◯ Anus<br>◯ Appendix<br>◯ Bile Ducts<br>◯ Bladder<br>◯ Brain<br>◯ Breast<br>◯ Cervical<br>◯ Colon (Large Intestine)<br>◯ Colorectal<br>◯ Esophagus<br>◯ Gallbladder<br>◯ Hodgkin's Lymphoma<br>◯ Kidney<br>◯ Leukemia<br>◯ Liver<br>◯ Lung<br>◯ Melanoma<br>◯ Mouth/throat<br>◯ Non-Hodgkin's Lymphoma<br>◯ Ovary<br>◯ Pancreas<br>◯ Pancreatic Islet Cell<br>◯ Parathyroid<br>◯ Pheochromocytoma<br>◯ Pituitary<br>◯ Prostate<br>◯ Rectum<br>◯ Small intestine<br>◯ Stomach<br>◯ Thyroid<br>◯ Uterine/Endometrial<br>◯ Unknown<br>◯ Other |
| What was his/her age at diagnosis? | __________________________ |

If more than one cancer, please include any additional cancer types and ages of diagnoses below. Please also elaborate is "other" was selected above as cancer type.

| | |
|---|---|
| Please select a different first-degree relative (please only describe FULL siblings, meaning siblings who share both biological parents with you): | ◯ Sibling 1<br>◯ Sibling 2<br>◯ Sibling 3<br>◯ Sibling 4<br>◯ Sibling 5<br>◯ Sibling 6<br>◯ Sibling 7<br>◯ Sibling 8<br>◯ Sibling 9<br>◯ Sibling 10<br>◯ Child 1<br>◯ Child 2<br>◯ Child 3<br>◯ Child 4<br>◯ Child 5<br>◯ Child 6<br>◯ Child 7<br>◯ Child 8<br>◯ Child 9<br>◯ Child 10<br>◯ I do not have anymore first degree relatives to describe |
| Sex: | ◯ Male<br>◯ Female |
| Is he/she still alive? | ◯ Yes<br>◯ No<br>◯ Unknown |
| What is his/her current age or at what age did he/she die? | ______________________________ |
| Has he/she ever had cancer? | ◯ Yes<br>◯ No<br>◯ Unknown |

| | |
|---|---|
| What type of cancer did he/she have? If more than one, include most recent cancer here, and see below. If other, please describe below. | ◯ Anus<br>◯ Appendix<br>◯ Bile Ducts<br>◯ Bladder<br>◯ Brain<br>◯ Breast<br>◯ Cervical<br>◯ Colon (Large Intestine)<br>◯ Colorectal<br>◯ Esophagus<br>◯ Gallbladder<br>◯ Hodgkin's Lymphoma<br>◯ Kidney<br>◯ Leukemia<br>◯ Liver<br>◯ Lung<br>◯ Melanoma<br>◯ Mouth/throat<br>◯ Non-Hodgkin's Lymphoma<br>◯ Ovary<br>◯ Pancreas<br>◯ Pancreatic Islet Cell<br>◯ Parathyroid<br>◯ Pheochromocytoma<br>◯ Pituitary<br>◯ Prostate<br>◯ Rectum<br>◯ Small intestine<br>◯ Stomach<br>◯ Thyroid<br>◯ Uterine/Endometrial<br>◯ Unknown<br>◯ Other |
| What was his/her age at diagnosis? | __________________________________ |

If more than one cancer, please include any additional cancer types and ages of diagnoses below. Please also elaborate if "other" was selected above as cancer type.

| | |
|---|---|
| Please select a different first-degree relative (please only describe FULL siblings, meaning siblings who share both biological parents with you): | ◯ Sibling 1<br>◯ Sibling 2<br>◯ Sibling 3<br>◯ Sibling 4<br>◯ Sibling 5<br>◯ Sibling 6<br>◯ Sibling 7<br>◯ Sibling 8<br>◯ Sibling 9<br>◯ Sibling 10<br>◯ Child 1<br>◯ Child 2<br>◯ Child 3<br>◯ Child 4<br>◯ Child 5<br>◯ Child 6<br>◯ Child 7<br>◯ Child 8<br>◯ Child 9<br>◯ Child 10<br>◯ I do not have anymore first degree relatives to describe |
| Sex: | ◯ Male<br>◯ Female |
| Is he/she still alive? | ◯ Yes<br>◯ No<br>◯ Unknown |
| What is his/her current age or at what age did he/she die? | __________________________________ |
| Has he/she ever had cancer? | ◯ Yes<br>◯ No<br>◯ Unknown |

What type of cancer did he/she have? If more than one, include most recent cancer here, and see below. If other, please describe below.

- ◯ Anus
- ◯ Appendix
- ◯ Bile Ducts
- ◯ Bladder
- ◯ Brain
- ◯ Breast
- ◯ Cervical
- ◯ Colon (Large Intestine)
- ◯ Colorectal
- ◯ Esophagus
- ◯ Gallbladder
- ◯ Hodgkin's Lymphoma
- ◯ Kidney
- ◯ Leukemia
- ◯ Liver
- ◯ Lung
- ◯ Melanoma
- ◯ Mouth/throat
- ◯ Non-Hodgkin's Lymphoma
- ◯ Ovary
- ◯ Pancreas
- ◯ Pancreatic Islet Cell
- ◯ Parathyroid
- ◯ Pheochromocytoma
- ◯ Pituitary
- ◯ Prostate
- ◯ Rectum
- ◯ Small intestine
- ◯ Stomach
- ◯ Thyroid
- ◯ Uterine/Endometrial
- ◯ Unknown
- ◯ Other

---

What was his/her age at diagnosis?

__________________________________

---

If more than one cancer, please include any additional cancer types and ages of diagnoses below. Please also elaborate if "other" was selected above as cancer type.

| | |
|---|---|
| Please select a different first-degree relative (please only describe FULL siblings, meaning siblings who share both biological parents with you): | ◯ Sibling 1<br>◯ Sibling 2<br>◯ Sibling 3<br>◯ Sibling 4<br>◯ Sibling 5<br>◯ Sibling 6<br>◯ Sibling 7<br>◯ Sibling 8<br>◯ Sibling 9<br>◯ Sibling 10<br>◯ Child 1<br>◯ Child 2<br>◯ Child 3<br>◯ Child 4<br>◯ Child 5<br>◯ Child 6<br>◯ Child 7<br>◯ Child 8<br>◯ Child 9<br>◯ Child 10<br>◯ I do not have anymore first degree relatives to describe |
| Sex: | ◯ Male<br>◯ Female |
| Is he/she still alive? | ◯ Yes<br>◯ No<br>◯ Unknown |
| What is his/her current age or at what age did he/she die? | ______________________ |
| Has he/she ever had cancer? | ◯ Yes<br>◯ No<br>◯ Unknown |

| | |
|---|---|
| What type of cancer did he/she have? If more than one, include most recent cancer here, and see below. If other, please describe below. | ◯ Anus<br>◯ Appendix<br>◯ Bile Ducts<br>◯ Bladder<br>◯ Brain<br>◯ Breast<br>◯ Cervical<br>◯ Colon (Large Intestine)<br>◯ Colorectal<br>◯ Esophagus<br>◯ Gallbladder<br>◯ Hodgkin's Lymphoma<br>◯ Kidney<br>◯ Leukemia<br>◯ Liver<br>◯ Lung<br>◯ Melanoma<br>◯ Mouth/throat<br>◯ Non-Hodgkin's Lymphoma<br>◯ Ovary<br>◯ Pancreas<br>◯ Pancreatic Islet Cell<br>◯ Parathyroid<br>◯ Pheochromocytoma<br>◯ Pituitary<br>◯ Prostate<br>◯ Rectum<br>◯ Small intestine<br>◯ Stomach<br>◯ Thyroid<br>◯ Uterine/Endometrial<br>◯ Unknown<br>◯ Other |
| What was his/her age at diagnosis? | ______________________________ |

If more than one cancer, please include any additional cancer types and ages of diagnoses below. Please also elaborate if "other" was selected above as cancer type.

| | |
|---|---|
| Please select a different first-degree relative (please only describe FULL siblings, meaning siblings who share both biological parents with you): | ◯ Sibling 1<br>◯ Sibling 2<br>◯ Sibling 3<br>◯ Sibling 4<br>◯ Sibling 5<br>◯ Sibling 6<br>◯ Sibling 7<br>◯ Sibling 8<br>◯ Sibling 9<br>◯ Sibling 10<br>◯ Child 1<br>◯ Child 2<br>◯ Child 3<br>◯ Child 4<br>◯ Child 5<br>◯ Child 6<br>◯ Child 7<br>◯ Child 8<br>◯ Child 9<br>◯ Child 10<br>◯ I do not have anymore first degree relatives to describe |
| Sex: | ◯ Male<br>◯ Female |
| Is he/she still alive? | ◯ Yes<br>◯ No<br>◯ Unknown |
| What is his/her current age or at what age did he/she die? | __________________________ |
| Has he/she ever had cancer? | ◯ Yes<br>◯ No<br>◯ Unknown |

What type of cancer did he/she have? If more than one, include most recent cancer here, and see below. If other, please describe below.

- ◯ Anus
- ◯ Appendix
- ◯ Bile Ducts
- ◯ Bladder
- ◯ Brain
- ◯ Breast
- ◯ Cervical
- ◯ Colon (Large Intestine)
- ◯ Colorectal
- ◯ Esophagus
- ◯ Gallbladder
- ◯ Hodgkin's Lymphoma
- ◯ Kidney
- ◯ Leukemia
- ◯ Liver
- ◯ Lung
- ◯ Melanoma
- ◯ Mouth/throat
- ◯ Non-Hodgkin's Lymphoma
- ◯ Ovary
- ◯ Pancreas
- ◯ Pancreatic Islet Cell
- ◯ Parathyroid
- ◯ Pheochromocytoma
- ◯ Pituitary
- ◯ Prostate
- ◯ Rectum
- ◯ Small intestine
- ◯ Stomach
- ◯ Thyroid
- ◯ Uterine/Endometrial
- ◯ Unknown
- ◯ Other

---

What was his/her age at diagnosis?

__________________________________

---

If more than one cancer, please include any additional cancer types and ages of diagnoses below. Please also elaborate if "other" was selected above as cancer type.

| | |
|---|---|
| Please select a different first-degree relative (please only describe FULL siblings, meaning siblings who share both biological parents with you): | ◯ Sibling 1<br>◯ Sibling 2<br>◯ Sibling 3<br>◯ Sibling 4<br>◯ Sibling 5<br>◯ Sibling 6<br>◯ Sibling 7<br>◯ Sibling 8<br>◯ Sibling 9<br>◯ Sibling 10<br>◯ Child 1<br>◯ Child 2<br>◯ Child 3<br>◯ Child 4<br>◯ Child 5<br>◯ Child 6<br>◯ Child 7<br>◯ Child 8<br>◯ Child 9<br>◯ Child 10<br>◯ I do not have anymore first degree relatives to describe |
| Sex: | ◯ Male<br>◯ Female |
| Is he/she still alive? | ◯ Yes<br>◯ No<br>◯ Unknown |
| What is his/her current age or at what age did he/she die? | __________________________________ |
| Has he/she ever had cancer? | ◯ Yes<br>◯ No<br>◯ Unknown |

What type of cancer did he/she have? If more than one, include most recent cancer here, and see below. If other, please describe below.

- ◯ Anus
- ◯ Appendix
- ◯ Bile Ducts
- ◯ Bladder
- ◯ Brain
- ◯ Breast
- ◯ Cervical
- ◯ Colon (Large Intestine)
- ◯ Colorectal
- ◯ Esophagus
- ◯ Gallbladder
- ◯ Hodgkin's Lymphoma
- ◯ Kidney
- ◯ Leukemia
- ◯ Liver
- ◯ Lung
- ◯ Melanoma
- ◯ Mouth/throat
- ◯ Non-Hodgkin's Lymphoma
- ◯ Ovary
- ◯ Pancreas
- ◯ Pancreatic Islet Cell
- ◯ Parathyroid
- ◯ Pheochromocytoma
- ◯ Pituitary
- ◯ Prostate
- ◯ Rectum
- ◯ Small intestine
- ◯ Stomach
- ◯ Thyroid
- ◯ Uterine/Endometrial
- ◯ Unknown
- ◯ Other

What was his/her age at diagnosis?

__________________________________

If more than one cancer, please include any additional cancer types and ages of diagnoses below. Please also elaborate if "other" was selected above as cancer type.

| | |
|---|---|
| Please select a different first-degree relative (please only describe FULL siblings, meaning siblings who share both biological parents with you): | ◯ Sibling 1<br>◯ Sibling 2<br>◯ Sibling 3<br>◯ Sibling 4<br>◯ Sibling 5<br>◯ Sibling 6<br>◯ Sibling 7<br>◯ Sibling 8<br>◯ Sibling 9<br>◯ Sibling 10<br>◯ Child 1<br>◯ Child 2<br>◯ Child 3<br>◯ Child 4<br>◯ Child 5<br>◯ Child 6<br>◯ Child 7<br>◯ Child 8<br>◯ Child 9<br>◯ Child 10<br>◯ I do not have anymore first degree relatives to describe |
| Sex: | ◯ Male<br>◯ Female |
| Is he/she still alive? | ◯ Yes<br>◯ No<br>◯ Unknown |
| What is his/her current age or at what age did he/she die? | __________________________ |
| Has he/she ever had cancer? | ◯ Yes<br>◯ No<br>◯ Unknown |

What type of cancer did he/she have? If more than one, include most recent cancer here, and see below. If other, please describe below.

- ◯ Anus
- ◯ Appendix
- ◯ Bile Ducts
- ◯ Bladder
- ◯ Brain
- ◯ Breast
- ◯ Cervical
- ◯ Colon (Large Intestine)
- ◯ Colorectal
- ◯ Esophagus
- ◯ Gallbladder
- ◯ Hodgkin's Lymphoma
- ◯ Kidney
- ◯ Leukemia
- ◯ Liver
- ◯ Lung
- ◯ Melanoma
- ◯ Mouth/throat
- ◯ Non-Hodgkin's Lymphoma
- ◯ Ovary
- ◯ Pancreas
- ◯ Pancreatic Islet Cell
- ◯ Parathyroid
- ◯ Pheochromocytoma
- ◯ Pituitary
- ◯ Prostate
- ◯ Rectum
- ◯ Small intestine
- ◯ Stomach
- ◯ Thyroid
- ◯ Uterine/Endometrial
- ◯ Unknown
- ◯ Other

---

What was his/her age at diagnosis?

__________________________________

---

If more than one cancer, please include any additional cancer types and ages of diagnoses below. Please also elaborate if "other" was selected above as cancer type.

| | |
|---|---|
| Please select a different first-degree relative (please only describe FULL siblings, meaning siblings who share both biological parents with you): | ◯ Sibling 1<br>◯ Sibling 2<br>◯ Sibling 3<br>◯ Sibling 4<br>◯ Sibling 5<br>◯ Sibling 6<br>◯ Sibling 7<br>◯ Sibling 8<br>◯ Sibling 9<br>◯ Sibling 10<br>◯ Child 1<br>◯ Child 2<br>◯ Child 3<br>◯ Child 4<br>◯ Child 5<br>◯ Child 6<br>◯ Child 7<br>◯ Child 8<br>◯ Child 9<br>◯ Child 10<br>◯ I do not have anymore first degree relatives to describe |
| Sex: | ◯ Male<br>◯ Female |
| Is he/she still alive? | ◯ Yes<br>◯ No<br>◯ Unknown |
| What is his/her current age or at what age did he/she die? | ______________________________ |
| Has he/she ever had cancer? | ◯ Yes<br>◯ No<br>◯ Unknown |

| | |
|---|---|
| What type of cancer did he/she have? If more than one, include most recent cancer here, and see below. If other, please describe below. | ◯ Anus<br>◯ Appendix<br>◯ Bile Ducts<br>◯ Bladder<br>◯ Brain<br>◯ Breast<br>◯ Cervical<br>◯ Colon (Large Intestine)<br>◯ Colorectal<br>◯ Esophagus<br>◯ Gallbladder<br>◯ Hodgkin's Lymphoma<br>◯ Kidney<br>◯ Leukemia<br>◯ Liver<br>◯ Lung<br>◯ Melanoma<br>◯ Mouth/throat<br>◯ Non-Hodgkin's Lymphoma<br>◯ Ovary<br>◯ Pancreas<br>◯ Pancreatic Islet Cell<br>◯ Parathyroid<br>◯ Pheochromocytoma<br>◯ Pituitary<br>◯ Prostate<br>◯ Rectum<br>◯ Small intestine<br>◯ Stomach<br>◯ Thyroid<br>◯ Uterine/Endometrial<br>◯ Unknown<br>◯ Other |
| What was his/her age at diagnosis? | ______________________________ |

If more than one cancer, please include any additional cancer types and ages of diagnoses below. Please also elaborate if "other" was selected above as cancer type.

| | |
|---|---|
| Please select a different first-degree relative (please only describe FULL siblings, meaning siblings who share both biological parents with you): | ◯ Sibling 1<br>◯ Sibling 2<br>◯ Sibling 3<br>◯ Sibling 4<br>◯ Sibling 5<br>◯ Sibling 6<br>◯ Sibling 7<br>◯ Sibling 8<br>◯ Sibling 9<br>◯ Sibling 10<br>◯ Child 1<br>◯ Child 2<br>◯ Child 3<br>◯ Child 4<br>◯ Child 5<br>◯ Child 6<br>◯ Child 7<br>◯ Child 8<br>◯ Child 9<br>◯ Child 10<br>◯ I do not have anymore first degree relatives to describe |
| Sex: | ◯ Male<br>◯ Female |
| Is he/she still alive? | ◯ Yes<br>◯ No<br>◯ Unknown |
| What is his/her current age or at what age did he/she die? | __________________________________ |
| Has he/she ever had cancer? | ◯ Yes<br>◯ No<br>◯ Unknown |

| | |
|---|---|
| What type of cancer did he/she have? If more than one, include most recent cancer here, and see below. If other, please describe below. | ◯ Anus<br>◯ Appendix<br>◯ Bile Ducts<br>◯ Bladder<br>◯ Brain<br>◯ Breast<br>◯ Cervical<br>◯ Colon (Large Intestine)<br>◯ Colorectal<br>◯ Esophagus<br>◯ Gallbladder<br>◯ Hodgkin's Lymphoma<br>◯ Kidney<br>◯ Leukemia<br>◯ Liver<br>◯ Lung<br>◯ Melanoma<br>◯ Mouth/throat<br>◯ Non-Hodgkin's Lymphoma<br>◯ Ovary<br>◯ Pancreas<br>◯ Pancreatic Islet Cell<br>◯ Parathyroid<br>◯ Pheochromocytoma<br>◯ Pituitary<br>◯ Prostate<br>◯ Rectum<br>◯ Small intestine<br>◯ Stomach<br>◯ Thyroid<br>◯ Uterine/Endometrial<br>◯ Unknown<br>◯ Other |
| What was his/her age at diagnosis? | __________________________ |

If more than one cancer, please include any additional cancer types and ages of diagnoses below. Please also elaborate is "other" was selected above as cancer type.

| | |
|---|---|
| Please select a different first-degree relative (please only describe FULL siblings, meaning siblings who share both biological parents with you): | ◯ Sibling 1<br>◯ Sibling 2<br>◯ Sibling 3<br>◯ Sibling 4<br>◯ Sibling 5<br>◯ Sibling 6<br>◯ Sibling 7<br>◯ Sibling 8<br>◯ Sibling 9<br>◯ Sibling 10<br>◯ Child 1<br>◯ Child 2<br>◯ Child 3<br>◯ Child 4<br>◯ Child 5<br>◯ Child 6<br>◯ Child 7<br>◯ Child 8<br>◯ Child 9<br>◯ Child 10<br>◯ I do not have anymore first degree relatives to describe |
| Sex: | ◯ Male<br>◯ Female |
| Is he/she still alive? | ◯ Yes<br>◯ No<br>◯ Unknown |
| What is his/her current age or at what age did he/she die? | ______________________________ |
| Has he/she ever had cancer? | ◯ Yes<br>◯ No<br>◯ Unknown |

What type of cancer did he/she have? If more than one, include most recent cancer here, and see below. If other, please describe below.

- ◯ Anus
- ◯ Appendix
- ◯ Bile Ducts
- ◯ Bladder
- ◯ Brain
- ◯ Breast
- ◯ Cervical
- ◯ Colon (Large Intestine)
- ◯ Colorectal
- ◯ Esophagus
- ◯ Gallbladder
- ◯ Hodgkin's Lymphoma
- ◯ Kidney
- ◯ Leukemia
- ◯ Liver
- ◯ Lung
- ◯ Melanoma
- ◯ Mouth/throat
- ◯ Non-Hodgkin's Lymphoma
- ◯ Ovary
- ◯ Pancreas
- ◯ Pancreatic Islet Cell
- ◯ Parathyroid
- ◯ Pheochromocytoma
- ◯ Pituitary
- ◯ Prostate
- ◯ Rectum
- ◯ Small intestine
- ◯ Stomach
- ◯ Thyroid
- ◯ Uterine/Endometrial
- ◯ Unknown
- ◯ Other

---

What was his/her age at diagnosis?

______________________________

---

If more than one cancer, please include any additional cancer types and ages of diagnoses below. Please also elaborate if "other" was selected above as cancer type.

| | |
|---|---|
| Please select a different first-degree relative (please only describe FULL siblings, meaning siblings who share both biological parents with you): | ◯ Sibling 1<br>◯ Sibling 2<br>◯ Sibling 3<br>◯ Sibling 4<br>◯ Sibling 5<br>◯ Sibling 6<br>◯ Sibling 7<br>◯ Sibling 8<br>◯ Sibling 9<br>◯ Sibling 10<br>◯ Child 1<br>◯ Child 2<br>◯ Child 3<br>◯ Child 4<br>◯ Child 5<br>◯ Child 6<br>◯ Child 7<br>◯ Child 8<br>◯ Child 9<br>◯ Child 10<br>◯ I do not have anymore first degree relatives to describe |
| Sex: | ◯ Male<br>◯ Female |
| Is he/she still alive? | ◯ Yes<br>◯ No<br>◯ Unknown |
| What is his/her current age or at what age did he/she die? | __________________________ |
| Has he/she ever had cancer? | ◯ Yes<br>◯ No<br>◯ Unknown |

| | |
|---|---|
| What type of cancer did he/she have? If more than one, include most recent cancer here, and see below. If other, please describe below. | ◯ Anus<br>◯ Appendix<br>◯ Bile Ducts<br>◯ Bladder<br>◯ Brain<br>◯ Breast<br>◯ Cervical<br>◯ Colon (Large Intestine)<br>◯ Colorectal<br>◯ Esophagus<br>◯ Gallbladder<br>◯ Hodgkin's Lymphoma<br>◯ Kidney<br>◯ Leukemia<br>◯ Liver<br>◯ Lung<br>◯ Melanoma<br>◯ Mouth/throat<br>◯ Non-Hodgkin's Lymphoma<br>◯ Ovary<br>◯ Pancreas<br>◯ Pancreatic Islet Cell<br>◯ Parathyroid<br>◯ Pheochromocytoma<br>◯ Pituitary<br>◯ Prostate<br>◯ Rectum<br>◯ Small intestine<br>◯ Stomach<br>◯ Thyroid<br>◯ Uterine/Endometrial<br>◯ Unknown<br>◯ Other |
| What was his/her age at diagnosis? | ______________________________ |

If more than one cancer, please include any additional cancer types and ages of diagnoses below. Please elaborate if "other" was selected above as cancer type.

| | |
|---|---|
| Please select a different first-degree relative (please only describe FULL siblings, meaning siblings who share both biological parents with you): | ◯ Sibling 1<br>◯ Sibling 2<br>◯ Sibling 3<br>◯ Sibling 4<br>◯ Sibling 5<br>◯ Sibling 6<br>◯ Sibling 7<br>◯ Sibling 8<br>◯ Sibling 9<br>◯ Sibling 10<br>◯ Child 1<br>◯ Child 2<br>◯ Child 3<br>◯ Child 4<br>◯ Child 5<br>◯ Child 6<br>◯ Child 7<br>◯ Child 8<br>◯ Child 9<br>◯ Child 10<br>◯ I do not have anymore first degree relatives to describe |
| Sex: | ◯ Male<br>◯ Female |
| Is he/she still alive? | ◯ Yes<br>◯ No<br>◯ Unknown |
| What is his/her current age or at what age did he/she die? | ______________________ |
| Has he/she ever had cancer? | ◯ Yes<br>◯ No<br>◯ Unknown |

| | |
|---|---|
| What type of cancer did he/she have? If more than one, include most recent cancer here, and see below. If other, please describe below. | ◯ Anus<br>◯ Appendix<br>◯ Bile Ducts<br>◯ Bladder<br>◯ Brain<br>◯ Breast<br>◯ Cervical<br>◯ Colon (Large Intestine)<br>◯ Colorectal<br>◯ Esophagus<br>◯ Gallbladder<br>◯ Hodgkin's Lymphoma<br>◯ Kidney<br>◯ Leukemia<br>◯ Liver<br>◯ Lung<br>◯ Melanoma<br>◯ Mouth/throat<br>◯ Non-Hodgkin's Lymphoma<br>◯ Ovary<br>◯ Pancreas<br>◯ Pancreatic Islet Cell<br>◯ Parathyroid<br>◯ Pheochromocytoma<br>◯ Pituitary<br>◯ Prostate<br>◯ Rectum<br>◯ Small intestine<br>◯ Stomach<br>◯ Thyroid<br>◯ Uterine/Endometrial<br>◯ Unknown<br>◯ Other |
| What was his/her age at diagnosis? | ______________________________ |

If more than one cancer, please include any additional cancer types and ages of diagnoses below. Please also elaborate if "other" was selected above as cancer type.

| | |
|---|---|
| Please select a different first-degree relative (please only describe FULL siblings, meaning siblings who share both biological parents with you): | ◯ Sibling 1<br>◯ Sibling 2<br>◯ Sibling 3<br>◯ Sibling 4<br>◯ Sibling 5<br>◯ Sibling 6<br>◯ Sibling 7<br>◯ Sibling 8<br>◯ Sibling 9<br>◯ Sibling 10<br>◯ Child 1<br>◯ Child 2<br>◯ Child 3<br>◯ Child 4<br>◯ Child 5<br>◯ Child 6<br>◯ Child 7<br>◯ Child 8<br>◯ Child 9<br>◯ Child 10<br>◯ I do not have anymore first degree relatives to describe |
| Sex: | ◯ Male<br>◯ Female |
| Is he/she still alive? | ◯ Yes<br>◯ No<br>◯ Unknown |
| What is his/her current age or at what age did he/she die? | __________________________________ |
| Has he/she ever had cancer? | ◯ Yes<br>◯ No<br>◯ Unknown |

What type of cancer did he/she have? If more than one, include most recent cancer here, and see below. If other, please describe below.

- ◯ Anus
- ◯ Appendix
- ◯ Bile Ducts
- ◯ Bladder
- ◯ Brain
- ◯ Breast
- ◯ Cervical
- ◯ Colon (Large Intestine)
- ◯ Colorectal
- ◯ Esophagus
- ◯ Gallbladder
- ◯ Hodgkin's Lymphoma
- ◯ Kidney
- ◯ Leukemia
- ◯ Liver
- ◯ Lung
- ◯ Melanoma
- ◯ Mouth/throat
- ◯ Non-Hodgkin's Lymphoma
- ◯ Ovary
- ◯ Pancreas
- ◯ Pancreatic Islet Cell
- ◯ Parathyroid
- ◯ Pheochromocytoma
- ◯ Pituitary
- ◯ Prostate
- ◯ Rectum
- ◯ Small intestine
- ◯ Stomach
- ◯ Thyroid
- ◯ Uterine/Endometrial
- ◯ Unknown
- ◯ Other

What was his/her age at diagnosis?

______________________________

If more than one cancer, please include any additional cancer types and ages of diagnoses below. Please also elaborate if "other" was selected above as cancer type.

| | |
|---|---|
| Please select a different first-degree relative (please only describe FULL siblings, meaning siblings who share both biological parents with you): | ◯ Sibling 1<br>◯ Sibling 2<br>◯ Sibling 3<br>◯ Sibling 4<br>◯ Sibling 5<br>◯ Sibling 6<br>◯ Sibling 7<br>◯ Sibling 8<br>◯ Sibling 9<br>◯ Sibling 10<br>◯ Child 1<br>◯ Child 2<br>◯ Child 3<br>◯ Child 4<br>◯ Child 5<br>◯ Child 6<br>◯ Child 7<br>◯ Child 8<br>◯ Child 9<br>◯ Child 10<br>◯ I do not have anymore first degree relatives to describe |
| Sex: | ◯ Male<br>◯ Female |
| Is he/she still alive? | ◯ Yes<br>◯ No<br>◯ Unknown |
| What is his/her current age or at what age did he/she die? | ______________________________ |
| Has he/she ever had cancer? | ◯ Yes<br>◯ No<br>◯ Unknown |

| | |
|---|---|
| What type of cancer did he/she have? If more than one, include most recent cancer here, and see below. If other, please describe below. | ◯ Anus<br>◯ Appendix<br>◯ Bile Ducts<br>◯ Bladder<br>◯ Brain<br>◯ Breast<br>◯ Cervical<br>◯ Colon (Large Intestine)<br>◯ Colorectal<br>◯ Esophagus<br>◯ Gallbladder<br>◯ Hodgkin's Lymphoma<br>◯ Kidney<br>◯ Leukemia<br>◯ Liver<br>◯ Lung<br>◯ Melanoma<br>◯ Mouth/throat<br>◯ Non-Hodgkin's Lymphoma<br>◯ Ovary<br>◯ Pancreas<br>◯ Pancreatic Islet Cell<br>◯ Parathyroid<br>◯ Pheochromocytoma<br>◯ Pituitary<br>◯ Prostate<br>◯ Rectum<br>◯ Small intestine<br>◯ Stomach<br>◯ Thyroid<br>◯ Uterine/Endometrial<br>◯ Unknown<br>◯ Other |
| What was his/her age at diagnosis? | ______________________________ |

If more than one cancer, please include any additional cancer types and ages of diagnoses below. Please also elaborate if "other" was selected above as cancer type.

| | |
|---|---|
| Please select a different first-degree relative (please only describe FULL siblings, meaning siblings who share both biological parents with you): | ◯ Sibling 1<br>◯ Sibling 2<br>◯ Sibling 3<br>◯ Sibling 4<br>◯ Sibling 5<br>◯ Sibling 6<br>◯ Sibling 7<br>◯ Sibling 8<br>◯ Sibling 9<br>◯ Sibling 10<br>◯ Child 1<br>◯ Child 2<br>◯ Child 3<br>◯ Child 4<br>◯ Child 5<br>◯ Child 6<br>◯ Child 7<br>◯ Child 8<br>◯ Child 9<br>◯ Child 10<br>◯ I do not have anymore first degree relatives to describe |
| Sex: | ◯ Male<br>◯ Female |
| Is he/she still alive? | ◯ Yes<br>◯ No<br>◯ Unknown |
| What is his/her current age or at what age did he/she die? | __________________________________ |
| Has he/she ever had cancer? | ◯ Yes<br>◯ No<br>◯ Unknown |

| | |
|---|---|
| What type of cancer did he/she have? If more than one, include most recent cancer here, and see below. If other, please describe below. | ◯ Anus<br>◯ Appendix<br>◯ Bile Ducts<br>◯ Bladder<br>◯ Brain<br>◯ Breast<br>◯ Cervical<br>◯ Colon (Large Intestine)<br>◯ Colorectal<br>◯ Esophagus<br>◯ Gallbladder<br>◯ Hodgkin's Lymphoma<br>◯ Kidney<br>◯ Leukemia<br>◯ Liver<br>◯ Lung<br>◯ Melanoma<br>◯ Mouth/throat<br>◯ Non-Hodgkin's Lymphoma<br>◯ Ovary<br>◯ Pancreas<br>◯ Pancreatic Islet Cell<br>◯ Parathyroid<br>◯ Pheochromocytoma<br>◯ Pituitary<br>◯ Prostate<br>◯ Rectum<br>◯ Small intestine<br>◯ Stomach<br>◯ Thyroid<br>◯ Uterine/Endometrial<br>◯ Unknown<br>◯ Other |
| What was his/her age at diagnosis? | ______________________________ |

If more than one cancer, please include any additional cancer types and ages of diagnoses below. Please also elaborate if "other" was selected above as cancer type.

| | |
|---|---|
| Please select a different first-degree relative (please only describe FULL siblings, meaning siblings who share both biological parents with you): | ◯ Sibling 1<br>◯ Sibling 2<br>◯ Sibling 3<br>◯ Sibling 4<br>◯ Sibling 5<br>◯ Sibling 6<br>◯ Sibling 7<br>◯ Sibling 8<br>◯ Sibling 9<br>◯ Sibling 10<br>◯ Child 1<br>◯ Child 2<br>◯ Child 3<br>◯ Child 4<br>◯ Child 5<br>◯ Child 6<br>◯ Child 7<br>◯ Child 8<br>◯ Child 9<br>◯ Child 10<br>◯ I do not have anymore first degree relatives to describe |
| Sex: | ◯ Male<br>◯ Female |
| Is he/she still alive? | ◯ Yes<br>◯ No<br>◯ Unknown |
| What is his/her current age or at what age did he/she die? | ______________________________ |
| Has he/she ever had cancer? | ◯ Yes<br>◯ No<br>◯ Unknown |

What type of cancer did he/she have? If more than one, include most recent cancer here, and see below. If other, please describe below.

- ◯ Anus
- ◯ Appendix
- ◯ Bile Ducts
- ◯ Bladder
- ◯ Brain
- ◯ Breast
- ◯ Cervical
- ◯ Colon (Large Intestine)
- ◯ Colorectal
- ◯ Esophagus
- ◯ Gallbladder
- ◯ Hodgkin's Lymphoma
- ◯ Kidney
- ◯ Leukemia
- ◯ Liver
- ◯ Lung
- ◯ Melanoma
- ◯ Mouth/throat
- ◯ Non-Hodgkin's Lymphoma
- ◯ Ovary
- ◯ Pancreas
- ◯ Pancreatic Islet Cell
- ◯ Parathyroid
- ◯ Pheochromocytoma
- ◯ Pituitary
- ◯ Prostate
- ◯ Rectum
- ◯ Small intestine
- ◯ Stomach
- ◯ Thyroid
- ◯ Uterine/Endometrial
- ◯ Unknown
- ◯ Other

---

What was his/her age at diagnosis?

__________________________________

---

If more than one cancer, please include any additional cancer types and ages of diagnoses below. Please elaborate if "other" was selected above as cancer type.

| | |
|---|---|
| Please select a different first-degree relative (please only describe FULL siblings, meaning siblings who share both biological parents with you): | ◯ Sibling 1<br>◯ Sibling 2<br>◯ Sibling 3<br>◯ Sibling 4<br>◯ Sibling 5<br>◯ Sibling 6<br>◯ Sibling 7<br>◯ Sibling 8<br>◯ Sibling 9<br>◯ Sibling 10<br>◯ Child 1<br>◯ Child 2<br>◯ Child 3<br>◯ Child 4<br>◯ Child 5<br>◯ Child 6<br>◯ Child 7<br>◯ Child 8<br>◯ Child 9<br>◯ Child 10<br>◯ I do not have anymore first degree relatives to describe |
| Sex: | ◯ Male<br>◯ Female |
| Is he/she still alive? | ◯ Yes<br>◯ No<br>◯ Unknown |
| What is his/her current age or at what age did he/she die? | ______________________________ |
| Has he/she ever had cancer? | ◯ Yes<br>◯ No<br>◯ Unknown |

| | |
|---|---|
| What type of cancer did he/she have? If more than one, include most recent cancer here, and see below. If other, please describe below. | ◯ Anus<br>◯ Appendix<br>◯ Bile Ducts<br>◯ Bladder<br>◯ Brain<br>◯ Breast<br>◯ Cervical<br>◯ Colon (Large Intestine)<br>◯ Colorectal<br>◯ Esophagus<br>◯ Gallbladder<br>◯ Hodgkin's Lymphoma<br>◯ Kidney<br>◯ Leukemia<br>◯ Liver<br>◯ Lung<br>◯ Melanoma<br>◯ Mouth/throat<br>◯ Non-Hodgkin's Lymphoma<br>◯ Ovary<br>◯ Pancreas<br>◯ Pancreatic Islet Cell<br>◯ Parathyroid<br>◯ Pheochromocytoma<br>◯ Pituitary<br>◯ Prostate<br>◯ Rectum<br>◯ Small intestine<br>◯ Stomach<br>◯ Thyroid<br>◯ Uterine/Endometrial<br>◯ Unknown<br>◯ Other |
| What was his/her age at diagnosis? | ______________________________ |

If more than one cancer, please include any additional cancer types and ages of diagnoses below. Please also elaborate if "other" was selected above as cancer type.

We are interested whether or not anyone else in your family has had any type of cancer. Please enter any second or third degree relatives (grandparents, grandchild, cousins, great aunt or uncle, nieces/nephews, half-siblings, etc) who have had cancer.

PLEASE ONLY DESCRIBE RELATIVES WHO HAVE HAD CANCER.

Select a relative from the initial drop down menu and give as much of the requested information as possible.

If you do not know a relative's exact age or age at diagnosis, please approximate as best you can. For example, type 65 if you think the relative is in his/her 60's.

| | | |
|---|---|---|
| 72 | Please select a second or third degree relative who has had cancer: | ◯ Grandparent<br>◯ Grandchild<br>◯ Uncle or Aunt<br>◯ Nephew or Niece<br>◯ Half-sibling (share only one biological parent)<br>◯ First Cousin (child of your parent's sibling)<br>◯ Great-Grandparent<br>◯ Great-Grandchild<br>◯ Great Uncle/Aunt (a sibling of your grandparent)<br>◯ Child of a Nephew of Niece<br>◯ Child of a Half-Sibling<br>◯ I have no second or third degree relatives who have been diagnosed with cancer |
| | Sex: | ◯ Male<br>◯ Female |
| | Which side of your family is he/she on? | ◯ Paternal<br>◯ Maternal |
| | Is he/she still alive? | ◯ Yes<br>◯ No<br>◯ Unknown |
| | What is his/her current age or at what age did he/she die? | __________________________ |

What type of cancer did he/she have? If more than one, include most recent cancer here and see below. If other, please describe below.

- ◯ Anus
- ◯ Appendix
- ◯ Bile Ducts
- ◯ Bladder
- ◯ Brain
- ◯ Breast
- ◯ Cervical
- ◯ Colon (Large Intestine)
- ◯ Colorectal
- ◯ Esophagus
- ◯ Gallbladder
- ◯ Hodgkin's Lymphoma
- ◯ Kidney
- ◯ Leukemia
- ◯ Liver
- ◯ Lung
- ◯ Melanoma
- ◯ Mouth/throat
- ◯ Non-Hodgkin's Lymphoma
- ◯ Ovary
- ◯ Pancreas
- ◯ Pancreatic Islet Cell
- ◯ Parathyroid
- ◯ Pheochromocytoma
- ◯ Pituitary
- ◯ Prostate
- ◯ Rectum
- ◯ Small intestine
- ◯ Stomach
- ◯ Thyroid
- ◯ Uterine/Endometrial
- ◯ Unknown
- ◯ Other

---

What was his/her age of diagnosis?

__________________________________

---

If more than one cancer, please include any additional cancer types and ages of diagnoses below. Please also elaborate if "other" was selected above as cancer type.

---

Please select a second or third degree relative who has had cancer:

- ◯ Grandparent
- ◯ Grandchild
- ◯ Uncle or Aunt
- ◯ Nephew or Niece
- ◯ Half-sibling (share only one biological parent)
- ◯ First Cousin (child of your parent's sibling)
- ◯ Great-Grandparent
- ◯ Great-Grandchild
- ◯ Great Uncle/Aunt (a sibling of your grandparent)
- ◯ Child of a Nephew of Niece
- ◯ Child of a Half-Sibling
- ◯ I have no more second or third degree relatives who have been diagnosed with cancer

---

Sex:

- ◯ Male
- ◯ Female

| | |
|---|---|
| Which side of your family is he/she on? | ◯ Paternal<br>◯ Maternal |
| Is he/she still alive? | ◯ Yes<br>◯ No<br>◯ Unknown |
| What is his/her current age or at what age did he/she die? | __________________________________ |
| What type of cancer did he/she have? If more than one, include most recent cancer here and see below. If other, please describe below. | ◯ Anus<br>◯ Appendix<br>◯ Bile Ducts<br>◯ Bladder<br>◯ Brain<br>◯ Breast<br>◯ Cervical<br>◯ Colon (Large Intestine)<br>◯ Colorectal<br>◯ Esophagus<br>◯ Gallbladder<br>◯ Hodgkin's Lymphoma<br>◯ Kidney<br>◯ Leukemia<br>◯ Liver<br>◯ Lung<br>◯ Melanoma<br>◯ Mouth/throat<br>◯ Non-Hodgkin's Lymphoma<br>◯ Ovary<br>◯ Pancreas<br>◯ Pancreatic Islet Cell<br>◯ Parathyroid<br>◯ Pheochromocytoma<br>◯ Pituitary<br>◯ Prostate<br>◯ Rectum<br>◯ Small intestine<br>◯ Stomach<br>◯ Thyroid<br>◯ Uterine/Endometrial<br>◯ Unknown<br>◯ Other |
| What was his/her age of diagnosis? | __________________________________ |

If more than one cancer, please include any additional cancer types and ages of diagnoses below. Please also elaborate if "other" was selected above as cancer type.

| | |
|---|---|
| Please select a second or third degree relative who has had cancer: | ◯ Grandparent<br>◯ Grandchild<br>◯ Uncle or Aunt<br>◯ Nephew or Niece<br>◯ Half-sibling (share only one biological parent)<br>◯ First Cousin (child of your parent's sibling)<br>◯ Great-Grandparent<br>◯ Great-Grandchild<br>◯ Great Uncle/Aunt (a sibling of your grandparent)<br>◯ Child of a Nephew of Niece<br>◯ Child of a Half-Sibling<br>◯ I have no more second or third degree relatives who have been diagnosed with cancer |
| Sex: | ◯ Male<br>◯ Female |
| Which side of your family is he/she on? | ◯ Paternal<br>◯ Maternal |
| Is he/she still alive? | ◯ Yes<br>◯ No<br>◯ Unknown |
| What is his/her current age or at what age did he/she die? | ______________________ |

| | |
|---|---|
| What type of cancer did he/she have? If more than one, include most recent cancer here and see below. If other, please describe below. | ◯ Anus<br>◯ Appendix<br>◯ Bile Ducts<br>◯ Bladder<br>◯ Brain<br>◯ Breast<br>◯ Cervical<br>◯ Colon (Large Intestine)<br>◯ Colorectal<br>◯ Esophagus<br>◯ Gallbladder<br>◯ Hodgkin's Lymphoma<br>◯ Kidney<br>◯ Leukemia<br>◯ Liver<br>◯ Lung<br>◯ Melanoma<br>◯ Mouth/throat<br>◯ Non-Hodgkin's Lymphoma<br>◯ Ovary<br>◯ Pancreas<br>◯ Pancreatic Islet Cell<br>◯ Parathyroid<br>◯ Pheochromocytoma<br>◯ Pituitary<br>◯ Prostate<br>◯ Rectum<br>◯ Small intestine<br>◯ Stomach<br>◯ Thyroid<br>◯ Uterine/Endometrial<br>◯ Unknown<br>◯ Other |
| What was his/her age of diagnosis? | ______________________________ |
| If more than one cancer, please include any additional cancer types and ages of diagnoses below. Please also elaborate if "other" was selected above as cancer type. | |
| Please select a second or third degree relative who has had cancer: | ◯ Grandparent<br>◯ Grandchild<br>◯ Uncle or Aunt<br>◯ Nephew or Niece<br>◯ Half-sibling (share only one biological parent)<br>◯ First Cousin (child of your parent's sibling)<br>◯ Great-Grandparent<br>◯ Great-Grandchild<br>◯ Great Uncle/Aunt (a sibling of your grandparent)<br>◯ Child of a Nephew of Niece<br>◯ Child of a Half-Sibling<br>◯ I have no more second or third degree relatives who have been diagnosed with cancer |
| Sex: | ◯ Male<br>◯ Female |

| | |
|---|---|
| Which side of your family is he/she on? | ◯ Paternal<br>◯ Maternal |
| Is he/she still alive? | ◯ Yes<br>◯ No<br>◯ Unknown |
| What is his/her current age or at what age did he/she die? | ______________________________ |
| What type of cancer did he/she have? If more than one, include most recent cancer here and see below. If other, please describe below. | ◯ Anus<br>◯ Appendix<br>◯ Bile Ducts<br>◯ Bladder<br>◯ Brain<br>◯ Breast<br>◯ Cervical<br>◯ Colon (Large Intestine)<br>◯ Colorectal<br>◯ Esophagus<br>◯ Gallbladder<br>◯ Hodgkin's Lymphoma<br>◯ Kidney<br>◯ Leukemia<br>◯ Liver<br>◯ Lung<br>◯ Melanoma<br>◯ Mouth/throat<br>◯ Non-Hodgkin's Lymphoma<br>◯ Ovary<br>◯ Pancreas<br>◯ Pancreatic Islet Cell<br>◯ Parathyroid<br>◯ Pheochromocytoma<br>◯ Pituitary<br>◯ Prostate<br>◯ Rectum<br>◯ Small intestine<br>◯ Stomach<br>◯ Thyroid<br>◯ Uterine/Endometrial<br>◯ Unknown<br>◯ Other |
| What was his/her age of diagnosis? | ______________________________ |

If more than one cancer, please include any additional cancer types and ages of diagnoses below. Please also elaborate if "other" was selected above as cancer type.

| | |
|---|---|
| Please select a second or third degree relative who has had cancer: | ◯ Grandparent<br>◯ Grandchild<br>◯ Uncle or Aunt<br>◯ Nephew or Niece<br>◯ Half-sibling (share only one biological parent)<br>◯ First Cousin (child of your parent's sibling)<br>◯ Great-Grandparent<br>◯ Great-Grandchild<br>◯ Great Uncle/Aunt (a sibling of your grandparent)<br>◯ Child of a Nephew of Niece<br>◯ Child of a Half-Sibling<br>◯ I have no more second or third degree relatives who have been diagnosed with cancer |
| Sex: | ◯ Male<br>◯ Female |
| Which side of your family is he/she on? | ◯ Paternal<br>◯ Maternal |
| Is he/she still alive? | ◯ Yes<br>◯ No<br>◯ Unknown |
| What is his/her current age or at what age did he/she die? | __________________________________ |

| | |
|---|---|
| What type of cancer did he/she have? If more than one, include most recent cancer here and see below. If other, please describe below. | ◯ Anus<br>◯ Appendix<br>◯ Bile Ducts<br>◯ Bladder<br>◯ Brain<br>◯ Breast<br>◯ Cervical<br>◯ Colon (Large Intestine)<br>◯ Colorectal<br>◯ Esophagus<br>◯ Gallbladder<br>◯ Hodgkin's Lymphoma<br>◯ Kidney<br>◯ Leukemia<br>◯ Liver<br>◯ Lung<br>◯ Melanoma<br>◯ Mouth/throat<br>◯ Non-Hodgkin's Lymphoma<br>◯ Ovary<br>◯ Pancreas<br>◯ Pancreatic Islet Cell<br>◯ Parathyroid<br>◯ Pheochromocytoma<br>◯ Pituitary<br>◯ Prostate<br>◯ Rectum<br>◯ Small intestine<br>◯ Stomach<br>◯ Thyroid<br>◯ Uterine/Endometrial<br>◯ Unknown<br>◯ Other |
| What was his/her age of diagnosis? | __________________________________ |

If more than one cancer, please include any additional cancer types and ages of diagnoses below. Please also elaborate if "other" was selected above as cancer type.

| | |
|---|---|
| Please select a second or third degree relative who has had cancer: | ◯ Grandparent<br>◯ Grandchild<br>◯ Uncle or Aunt<br>◯ Nephew or Niece<br>◯ Half-sibling (share only one biological parent)<br>◯ First Cousin (child of your parent's sibling)<br>◯ Great-Grandparent<br>◯ Great-Grandchild<br>◯ Great Uncle/Aunt (a sibling of your grandparent)<br>◯ Child of a Nephew of Niece<br>◯ Child of a Half-Sibling<br>◯ I have no more second or third degree relatives who have been diagnosed with cancer |
| Sex: | ◯ Male<br>◯ Female |

| | |
|---|---|
| Which side of your family is he/she on? | ◯ Paternal<br>◯ Maternal |
| Is he/she still alive? | ◯ Yes<br>◯ No<br>◯ Unknown |
| What is his/her current age or at what age did he/she die? | __________________________________ |
| What type of cancer did he/she have? If more than one, include most recent cancer here and see below. If other, please describe below. | ◯ Anus<br>◯ Appendix<br>◯ Bile Ducts<br>◯ Bladder<br>◯ Brain<br>◯ Breast<br>◯ Cervical<br>◯ Colon (Large Intestine)<br>◯ Colorectal<br>◯ Esophagus<br>◯ Gallbladder<br>◯ Hodgkin's Lymphoma<br>◯ Kidney<br>◯ Leukemia<br>◯ Liver<br>◯ Lung<br>◯ Melanoma<br>◯ Mouth/throat<br>◯ Non-Hodgkin's Lymphoma<br>◯ Ovary<br>◯ Pancreas<br>◯ Pancreatic Islet Cell<br>◯ Parathyroid<br>◯ Pheochromocytoma<br>◯ Pituitary<br>◯ Prostate<br>◯ Rectum<br>◯ Small intestine<br>◯ Stomach<br>◯ Thyroid<br>◯ Uterine/Endometrial<br>◯ Unknown<br>◯ Other |
| What was his/her age of diagnosis? | __________________________________ |

If more than one cancer, please include any additional cancer types and ages of diagnoses below. Please also elaborate if "other" was selected above as cancer type.

| | |
|---|---|
| Please select a second or third degree relative who has had cancer: | ◯ Grandparent<br>◯ Grandchild<br>◯ Uncle or Aunt<br>◯ Nephew or Niece<br>◯ Half-sibling (share only one biological parent)<br>◯ First Cousin (child of your parent's sibling)<br>◯ Great-Grandparent<br>◯ Great-Grandchild<br>◯ Great Uncle/Aunt (a sibling of your grandparent)<br>◯ Child of a Nephew of Niece<br>◯ Child of a Half-Sibling<br>◯ I have no more second or third degree relatives who have been diagnosed with cancer |
| Sex: | ◯ Male<br>◯ Female |
| Which side of your family is he/she on? | ◯ Paternal<br>◯ Maternal |
| Is he/she still alive? | ◯ Yes<br>◯ No<br>◯ Unknown |
| What is his/her current age or at what age did he/she die? | __________________________ |

What type of cancer did he/she have? If more than one, include most recent cancer here and see below. If other, please describe below.

- ◯ Anus
- ◯ Appendix
- ◯ Bile Ducts
- ◯ Bladder
- ◯ Brain
- ◯ Breast
- ◯ Cervical
- ◯ Colon (Large Intestine)
- ◯ Colorectal
- ◯ Esophagus
- ◯ Gallbladder
- ◯ Hodgkin's Lymphoma
- ◯ Kidney
- ◯ Leukemia
- ◯ Liver
- ◯ Lung
- ◯ Melanoma
- ◯ Mouth/throat
- ◯ Non-Hodgkin's Lymphoma
- ◯ Ovary
- ◯ Pancreas
- ◯ Pancreatic Islet Cell
- ◯ Parathyroid
- ◯ Pheochromocytoma
- ◯ Pituitary
- ◯ Prostate
- ◯ Rectum
- ◯ Small intestine
- ◯ Stomach
- ◯ Thyroid
- ◯ Uterine/Endometrial
- ◯ Unknown
- ◯ Other

---

What was his/her age of diagnosis?

______________________________

---

If more than one cancer, please include any additional cancer types and ages of diagnoses below. Please also elaborate if "other" was selected above as cancer type.

---

Please select a second or third degree relative who has had cancer:

- ◯ Grandparent
- ◯ Grandchild
- ◯ Uncle or Aunt
- ◯ Nephew or Niece
- ◯ Half-sibling (share only one biological parent)
- ◯ First Cousin (child of your parent's sibling)
- ◯ Great-Grandparent
- ◯ Great-Grandchild
- ◯ Great Uncle/Aunt (a sibling of your grandparent)
- ◯ Child of a Nephew of Niece
- ◯ Child of a Half-Sibling
- ◯ I have no more second or third degree relatives who have been diagnosed with cancer

---

Sex:

- ◯ Male
- ◯ Female

| | |
|---|---|
| Which side of your family is he/she on? | ◯ Paternal<br>◯ Maternal |
| Is he/she still alive? | ◯ Yes<br>◯ No<br>◯ Unknown |
| What is his/her current age or at what age did he/she die? | __________________________________ |
| What type of cancer did he/she have? If more than one, include most recent cancer here and see below. If other, please describe below. | ◯ Anus<br>◯ Appendix<br>◯ Bile Ducts<br>◯ Bladder<br>◯ Brain<br>◯ Breast<br>◯ Cervical<br>◯ Colon (Large Intestine)<br>◯ Colorectal<br>◯ Esophagus<br>◯ Gallbladder<br>◯ Hodgkin's Lymphoma<br>◯ Kidney<br>◯ Leukemia<br>◯ Liver<br>◯ Lung<br>◯ Melanoma<br>◯ Mouth/throat<br>◯ Non-Hodgkin's Lymphoma<br>◯ Ovary<br>◯ Pancreas<br>◯ Pancreatic Islet Cell<br>◯ Parathyroid<br>◯ Pheochromocytoma<br>◯ Pituitary<br>◯ Prostate<br>◯ Rectum<br>◯ Small intestine<br>◯ Stomach<br>◯ Thyroid<br>◯ Uterine/Endometrial<br>◯ Unknown<br>◯ Other |
| What was his/her age of diagnosis? | __________________________________ |

If more than one cancer, please include any additional cancer types and ages of diagnoses below. Please also elaborate if "other" was selected above as cancer type.

| | |
|---|---|
| Please select a second or third degree relative who has had cancer: | ◯ Grandparent<br>◯ Grandchild<br>◯ Uncle or Aunt<br>◯ Nephew or Niece<br>◯ Half-sibling (share only one biological parent)<br>◯ First Cousin (child of your parent's sibling)<br>◯ Great-Grandparent<br>◯ Great-Grandchild<br>◯ Great Uncle/Aunt (a sibling of your grandparent)<br>◯ Child of a Nephew of Niece<br>◯ Child of a Half-Sibling<br>◯ I have no more second or third degree relatives who have been diagnosed with cancer |
| Sex: | ◯ Male<br>◯ Female |
| Which side of your family is he/she on? | ◯ Paternal<br>◯ Maternal |
| Is he/she still alive? | ◯ Yes<br>◯ No<br>◯ Unknown |
| What is his/her current age or at what age did he/she die? | ____________________ |

What type of cancer did he/she have? If more than one, include most recent cancer here and see below. If other, please describe below.

- ◯ Anus
- ◯ Appendix
- ◯ Bile Ducts
- ◯ Bladder
- ◯ Brain
- ◯ Breast
- ◯ Cervical
- ◯ Colon (Large Intestine)
- ◯ Colorectal
- ◯ Esophagus
- ◯ Gallbladder
- ◯ Hodgkin's Lymphoma
- ◯ Kidney
- ◯ Leukemia
- ◯ Liver
- ◯ Lung
- ◯ Melanoma
- ◯ Mouth/throat
- ◯ Non-Hodgkin's Lymphoma
- ◯ Ovary
- ◯ Pancreas
- ◯ Pancreatic Islet Cell
- ◯ Parathyroid
- ◯ Pheochromocytoma
- ◯ Pituitary
- ◯ Prostate
- ◯ Rectum
- ◯ Small intestine
- ◯ Stomach
- ◯ Thyroid
- ◯ Uterine/Endometrial
- ◯ Unknown
- ◯ Other

---

What was his/her age of diagnosis?

______________________________

---

If more than one cancer, please include any additional cancer types and ages of diagnoses below. Please also elaborate if "other" was selected above as cancer type.

---

Please select a second or third degree relative who has had cancer:

- ◯ Grandparent
- ◯ Grandchild
- ◯ Uncle or Aunt
- ◯ Nephew or Niece
- ◯ Half-sibling (share only one biological parent)
- ◯ First Cousin (child of your parent's sibling)
- ◯ Great-Grandparent
- ◯ Great-Grandchild
- ◯ Great Uncle/Aunt (a sibling of your grandparent)
- ◯ Child of a Nephew of Niece
- ◯ Child of a Half-Sibling
- ◯ I have no more second or third degree relatives who have been diagnosed with cancer

---

Sex:

- ◯ Male
- ◯ Female

| | |
|---|---|
| Which side of your family is he/she on? | ◯ Paternal<br>◯ Maternal |
| Is he/she still alive? | ◯ Yes<br>◯ No<br>◯ Unknown |
| What is his/her current age or at what age did he/she die? | __________________________________ |
| What type of cancer did he/she have? If more than one, include most recent cancer here and see below. If other, please describe below. | ◯ Anus<br>◯ Appendix<br>◯ Bile Ducts<br>◯ Bladder<br>◯ Brain<br>◯ Breast<br>◯ Cervical<br>◯ Colon (Large Intestine)<br>◯ Colorectal<br>◯ Esophagus<br>◯ Gallbladder<br>◯ Hodgkin's Lymphoma<br>◯ Kidney<br>◯ Leukemia<br>◯ Liver<br>◯ Lung<br>◯ Melanoma<br>◯ Mouth/throat<br>◯ Non-Hodgkin's Lymphoma<br>◯ Ovary<br>◯ Pancreas<br>◯ Pancreatic Islet Cell<br>◯ Parathyroid<br>◯ Pheochromocytoma<br>◯ Pituitary<br>◯ Prostate<br>◯ Rectum<br>◯ Small intestine<br>◯ Stomach<br>◯ Thyroid<br>◯ Uterine/Endometrial<br>◯ Unknown<br>◯ Other |
| What was his/her age of diagnosis? | __________________________________ |

If more than one cancer, please include any additional cancer types and ages of diagnoses below. Please also elaborate if "other" was selected above as cancer type.

73 If you ran out of space above, please describe any additional siblings, children, second, or third degree relatives here.

#### Extra Information

74 Please feel free to add any other information that you feel might be useful to the research on gastric cancer or you may use this space to clarify any answers on the questionnaire (please note question number).  Thank you.
