## Supplemental File 2 for "The Gastric Cancer Registry: A Genomic Translational Resource for Multidisciplinary Research in Stomach Malignancies"

**SUPPLEMENTARY FILE 2**

### TITLE

The Gastric Cancer Registry: A Genomic Translational Resource for Multidisciplinary Research in Stomach Malignancies


### AUTHORS

Alison F Almeda[1], Sue Grimes[1], HoJoon Lee[1], Stephanie U Greer[1], GiWon Shin[1], Madeline McNamara[1], Anna C Hooker[1], Maya Arce[1], Matthew Kubit[1], Marie Schauer[1], Paul Van Hummelen[1], Cindy Ma[1], Meredith Mills[1], Robert J Huang[2], Joo Ha Hwang[2], Manuel R Amieva[3], Summer Han[4], James M. Ford[1], Hanlee P. Ji[1*]

[1]Division of Oncology, Department of Medicine, Stanford University School of Medicine, Stanford, CA, 94305, United States

[2]Division of Gastroenterology and Hepatology, Department of Medicine, Stanford University School of Medicine, Stanford, CA, 94305, United States

[3]Division of Infectious Diseases, Department of Pediatrics, Stanford University School of Medicine, Stanford, CA, 94305, United States

[4]Department of Neurosurgery, Stanford University School of Medicine, Stanford, CA, 94305, United States

### CORRESPONDING AUTHOR

Hanlee P. Ji

Division of Oncology, Department of Medicine – Stanford University School of Medicine

CCSR 2245, 269 Campus Drive

Stanford, CA 94305-5151

### MATERIALS AND METHODS

#### DNA and RNA isolation

Scrolls measuring 5-20 μm were generated from FFPE blocks. Genomic DNA and total RNA were extracted using the Maxwell® 16 system (Promega Corporation, catalog no. AS1130, PRAS1260). Nucleic acids were quantified using the Qubit 2.0 Fluorometer (Thermo Fisher Scientific, catalog no. Q32866, Q10211). The quality of genomic DNA and RNA was assessed in terms of Genomic Quality Score and RNA Quality Score using the LabChip GX HT Touch Nucleic Acid Analyzer (PerkinElmer, catalog no. CLS137031).

#### Sequencing

For sequencing library preparation, 500 ng of DNA of each sample was sheared using a Covaris E220 (Covaris, Inc., catalog no. E220) with AFA microtubes (Covaris, Inc., catalog no. 520166) and parameters as follows: intensity level of five, duty cycle of 10%, cycles per burst of 200, and treatment time of 55 seconds. The DNA was then purified with a 0.8X AMPure XP (Beckman Coulter, Inc., catalog no. A63882) bead cleanup to maintain a higher DNA insert size for sequencing. The total yield of purified DNA was used as input for the KAPA Hyper Prep Kit for Illumina (Roche, catalog no. KK8502). The standard protocol was followed with 8 cycles of PCR amplification and a modification of a 0.8X post-amplification cleanup instead of the recommended 1.0X. The sequencing adapters used included 8 base pair unique dual indexes to allow for index swapping detection. A portion of the genomic libraries were enriched for exons using xGen Lockdown Probes and reagents (Integrated DNA Technologies, catalog no. 1056114, 1075474).

For RNA sequencing **(RNA-seq)**, the KAPA RNA HyperPrep Kit (Roche, catalog no. KK8540) was used to prepare libraries as per the manufacturer's protocol with some revisions. The adapter ligation time was extended to 1 hour and followed by a two-step bead clean up using KAPA Pure Beads (Roche, catalog no. KR1245). Libraries underwent a minimum of 15 PCR amplification cycles and were eluted with an additional 10□L elution buffer to account for samples with low quality RNA.

The whole genome, whole exome, and RNA-seq libraries were pooled based on the library type, quantified using real time PCR and then run on the iSeq 100 system (Illumina, catalog no. 20021532) for paired-end 150 base pair sequencing. Based upon the iSeq data, the sequencing libraries were re-pooled and normalized for higher depth 150 base paired end sequencing with a NovaSeq 6000 system (Illumina, catalog no. 20012850).

Genomic DNA libraries were sequenced for low-depth whole genome coverage between 1-2X. Paired-end reads were aligned to GRCh38 with the BWA-MEM algorithm (v0.7.15-r1140). Duplicate reads were marked using the Sentieon (v201711.04) LocusCollector algorithm. Copy number segments were called with CNVkit (v0.9.6.dev0). To identify somatic copy number changes for samples without a matched normal control, we used a normal reference genome data set as a comparison control.[1]

Exome libraries were sequenced at an average depth of 68X coverage (median depth of 58X coverage). The paired-end reads were aligned to GRCh38 with the BWA-MEM algorithm (v0.7.15-r1140). Duplicate reads were marked using the Sentieon (v201711.04) LocusCollector algorithm. GATK best practices were followed to call single nucleotide variants (SNVs) and indels using Sentieon (v201808.07). Variant calling involved first realigning intervals using the ReAligner Program and recalibrating the base quality score using QualCal. Variant calls were then made using TNhaplotyper2 in tumor-only mode, with the publicly available panel of normals: gatk4_mutect2_4136_pon.vcf (https://gdc.cancer.gov/about-data/gdc-data-processing/gdc-reference-files). Modeling and filtering for FFPE artifacts and cross-sample contamination was performed using the OrietnationBias and ContaminationModel algorithms respectively. Finally, VarCal and ApplyVarCal were used to recalibrate the single nucleotide variant and insertion / deletion variant quality score.

RNA libraries were sequenced at high depth for an average of 65 million reads per sample. The paired-end reads were aligned to GRCh38 by a two-pass method with STAR.[2] Gene expression level was measured in fragment of kb of transcript per million mapped reads using HT-Seq (v0.5.4p5). The four-digit human leukocyte antigen **(HLA)** genotypes of each sample were identified using OptiType.[3] Cellular deconvolution and quantitation of the abundance of 22 cell types was determined with the program CIBERSORTx, to identify tumor immune infiltrates. To characterize the microbiome of each sample, we used the Kraken2 application.[4] Unmapped reads from RNA-seq were extracted and mapped back to a Kraken2 database (version 2019.09). This database contained the complete genomes for human, bacterial, archaeal, and viral domains, as well as the taxonomic information from NCBI.

We identified candidate cancer neoantigens based on the following criteria: i) non-synonymous mutations, or amino acid changes, ii) expression in RNA-seq, and iii) binding to one of the patient's HLAs. First, we identified expressed nonsynonymous mutations from whole exome sequencing which were also expressed based on RNA-seq. Second, the sequences around the cancer mutations were extracted and translated into epitopes of nine amino acids long. We used the Consensus coding sequence (CCDS) database to define the coding sequence. This resource provides a comprehensive list of curated protein sequences derived from multiple international databases.[5] Finally, we used the program NetMHCpan 4.0 to predict the binding affinity of mutant epitopes to the HLA types per a given sample predicted by OptiType.[6] Based on the Rank%, which is a percentile score from NetMHCpan4.0, the epitopes a Rank% less than 0.5 were considered strong binders while ones with a Rank% between 0.5 and 2.0 were considered weak binders.
